# Adverse childhood experiences and multimorbidity of internalising and cardiometabolic conditions in mid to older age

**DOI:** 10.1101/2025.10.27.25338876

**Authors:** Lauren K. Benger, Peter Holmans, Michael J. Owen, Mark Mon-Williams, Frances Rice, Rupert Payne, Ioanna K. Katzourou, LINC Consortium, Marianne B.M. van den Bree

## Abstract

**Background:** Multimorbidity of internalising and cardiometabolic conditions (ICM-MM) is the most common combination of mental and physical health conditions in older age. Few studies have examined the likelihood that individuals with adverse childhood experiences (ACEs) such as abuse or neglect will develop ICM-MM in mid to late adulthood, or gender disparities.

**Methods:** UK Biobank participants (n = 157,184, mean age 55.94, SD= 7.74; 68186 males and 88998 females) reported on ACEs as well as sociodemographic and lifestyle factors. Diagnoses of internalising conditions (depression and anxiety) and cardiometabolic conditions (hypertension, obesity, type 2 diabetes, dyslipidaemia and chronic kidney disease) were obtained through linked electronic healthcare records. Logistic regression models tested associations between ACEs and internalising conditions, cardiometabolic conditions and ICM-MM, accounting for gender differences and sociodemographic and lifestyle factors.

**Results:** ACEs were associated with all individual and multimorbid presentations. Stronger associations were found with internalising conditions (OR 1.84) and ICM-MM (OR range 1.73-2.15) than cardiometabolic conditions (OR range 1.08-1.44). Females were more likely to report most ACEs, but health risks following ACEs were similar for both genders. The associations remained when accounting for sociodemographic and lifestyle factors, including gender, age, socioeconomic status, ethnicity, diet, alcohol intake, smoking status and physical activity levels.

**Conclusions:** This is the first study to report associations between ACEs and the most common type of physical and mental health multimorbidity in mid to late adulthood. The results highlight the importance of early ACE intervention and trauma-informed healthcare.

## Introduction

Multimorbidity refers to individuals experiencing two or more chronic conditions at the same time and has been identified as a major and increasing challenge to health care systems (Academy of Medical Sciences, 2018). It is particularly prevalent in the older population, being present in more than half of those aged over 60 years (Carlson & Yarns, 2023; Chowdhury, Das, Sunna, Beyene, & Hossain, 2023).

The most common physical and mental health multimorbidity cluster in older age is between internalising conditions (IC), such as anxiety or depression, and cardiometabolic conditions (CMC; preventable chronic diseases impacting the cardiovascular system and metabolic health). We will refer to this cluster of conditions as ICM-MM from here onwards. ICM-MM is associated with high levels of disability, increased complexity of disease management (S. W. Mercer, Gunn, Bower, Wyke, & Guthrie, 2012), high treatment burden for patients, poorer delivery of healthcare across fragmented health services (S. Mercer, Furler, Moffat, Fischbacher-Smith, & Sanci, 2016) and high healthcare costs (Soley-Bori et al., 2021; Stokes, Guthrie, Mercer, Rice, & Sutton, 2021).

CMCs such as type 2 diabetes (T2D), obesity, hypertension, chronic kidney disease (CKD) and dyslipidaemia are seen as precursors to cardiovascular disease and occur earlier in adulthood. Moreover, research indicates that risk of both IC and CMC start early in life, often before adulthood (Beesdo, Knappe, & Pine, 2009; Berenson et al., 1989; McGill Jr et al., 2000).

There is therefore a pressing need to understand the early life risk factors contributing to the development of ICM-MM in mid to late adulthood, as these insights may point to opportunities for prevention or early intervention.

Adverse childhood experiences (ACEs), such as abuse and neglect, have been associated with a wide range of mental and physical health conditions in adulthood (Felitti et al., 1998). Specifically, ACEs are associated with increased likelihood of ICs (Merrick et al., 2017) and CMCs (Suglia et al., 2018), including T2D (Hughes, Ford, & Bellis, 2020), obesity (Blissett, 2011), dyslipidaemia (Lin, Wang, Lu, Chen, & Guo, 2021), hypertension (Obi, McPherson, & Pollock, 2019) and CKD (Shi, Huang, & Jin, 2022). There are numerous studies on the association between ACEs and individual health outcomes but their impact on multimorbidity development in older age remains poorly understood, particularly for combinations of physical and mental health conditions. Exceptions are studies by Taylor and Demakakos (2024) and Hanlon et al. (2020), which have reported that experiencing a greater number of ACEs increases the risk of multimorbidity of mental and physical health conditions in older age, including ICs, and certain CMCs (i.e., diabetes, hypertension and CKD). However, these studies are based on self-reported conditions, which are liable to misreporting due to recall bias and limited health literacy (Sulieman et al., 2022) and need to be replicated using routinely recorded diagnoses from electronic health records (EHR). These studies also used counts of long-term conditions or disease categories, rather than combinations of specific mental and physical health conditions, to investigate multimorbidity. Identifying the precise conditions associated with ACEs would provide more meaningful information for both patients and clinicians, thereby facilitating the development of more targeted interventions.

There are well-documented gender differences in ACEs indicating that females are more likely to experience a higher frequency of ACEs and more complex ACE patterns than males (Haahr-Pedersen et al., 2020). Females are also more likely to experience most types of ACE, with the exception of physical abuse which has a similar or higher prevalence in males (Office for National Statistics, 2020a, 2020b). Furthermore, the risk of IC and CMC also tends to differ between the genders, with depression and anxiety more common in females (McLean, Asnaani, Litz, & Hofmann, 2011; Piccinelli & Wilkinson, 2000), whilst the prevalence of diagnosis of T2D, obesity, hypertension, dyslipidaemia and CKD differs between the genders in an age-dependent manner (Meloni et al., 2023; The World Health Organisation, 2023a, 2023b). It is therefore important that research into the associations between ACEs and ICM-MM examines gender differences.

Finally, a number of demographic and lifestyle factors, including lower socioeconomic status (SES), ethnic minority status, poor diet, smoking, high levels of alcohol use, and a sedentary lifestyle, are likely to influence the associations between ACEs, ICs and CMCs (Mersky, Choi, Lee, & Janczewski, 2021; Walsh, McCartney, Smith, & Armour, 2019). However, previous studies have not fully account for these factors (Hanlon et al., 2020; Taylor & Demakakos, 2024). Taking these factors into account will allow for better understanding of the relationships between ACES and individual condition and multimorbid presentations.

Our aims therefore were to examine:

1. Whether individuals in mid to older age who have experienced a range of ACEs (e.g., emotional, physical and sexual abuse, and emotional and physical neglect) are more likely to develop IC, CMC or ICM-MM than those without such experiences;
2. To what extent the associations between ACEs and IC, CMC or ICM-MM are impacted by demographic and lifestyle factors;
3. Whether the associations between ACEs and IC, CMC or ICM-MM differ between females and males.

## Methods

### Sample

Data were collated from the UK Biobank cohort (Bycroft et al., 2018). The cohort contains information on around 500,000 people in the UK who were aged between 38-72 years when they were recruited between 2006 and 2010. Data were released under application number 79704 “Physical and mental health multimorbidity across the lifespan (LIfespaN multimorbidity research Collaborative: LINC)”. This work included participants with data on at least one ACE as well as diagnoses of IC and CMC from EHR data (n = 157,184).

### Measures

#### Adverse Childhood Experiences

Following recruitment, 157,305 participants completed an online follow-up mental health questionnaire (MHQ), which queried ACEs, with responses from 157,184 participants used in analyses (see *Supplementary* Figure 1). The Childhood Trauma Screener (Grabe et al., 2012), was used to assess five ACE domains: emotional neglect, physical abuse, emotional abuse, sexual abuse and physical neglect. A binary variable was derived to indicate the presence of each ACE type following previous practice (Ho et al., 2020) (see *Supplementary Table 1* for details and coding of ACEs). Responses of “Prefer not to say” were coded as missing data. Those who gave a response to at least one of the five questions were included in the analysis but anyone who answered “Prefer not to say” to all five questions was removed (n = 42) (see *Supplementary* Figure 1).

#### Internalising Conditions and Cardiometabolic Conditions

Diagnoses of ICs and CMCs were available through linked EHRs for both primary (GP records) and secondary care (emergency and non-emergency hospital admission records). Primary care records are available for 45% of the UK Biobank cohort (Read v2 and CTV3 coding systems) and secondary care records for the full cohort (ICD-9 codes and ICD-10) (UK Biobank, 2024). In line with Katzourou et al. (2025), we combined the information on the presence or absence of depression and/or anxiety disorder to derive the variable “any IC”, because the two conditions tend to co-occur and respond to similar pharmacological and psychological treatments (Garber et al., 2016; Goodwin & Stein, 2021). To measure CMCs, diagnoses of hypertension, T2D, obesity, CKD and/or dyslipidaemia from

EHRs were used in line with (Katzourou et al., *in prep*)). A variable “any CMC” was derived to indicate the presence of one or more of the CMCs. Pairwise combinations of the presence of IC with each CMC individually as well as with any CMC were derived to measure ICM-MM. There were therefore eight derived outcome variables: any IC, any CMC, pairwise combinations of diagnoses of IC plus CMC (any IC plus T2D, any IC plus hypertension, any IC plus obesity, any IC plus CKD, any IC plus dyslipidaemia); and any ICM-MM (see *Supplementary Table 2* for coding of derived variables). These groupings are not mutually exclusive.

#### Demographic variables

Gender (female and male), age, SES and ethnicity, all collected at recruitment, were included in analyses. SES was measured using the Townsend Deprivation Index (TDI). A score of zero indicates the average material value of an area, positive values indicate high material deprivation and negative values indicate relative affluence. Ethnicity was categorised as white and non-white.

#### Lifestyle variables

Lifestyle variables (alcohol intake, diet, smoking status and physical activity) were taken from the touchscreen questionnaire completed at recruitment. Alcohol consumption was self-reported and coded as units per week (with >100 considered as missing data). A healthy diet score (ranging from ‘0’ least healthy to ‘7’ most healthy) was derived using data collected on consumption of 7 food groups (Hepsomali & Groeger, 2021; Wang et al., 2022). Smoker status was defined as never, previous or current. Physical activity was determined as total Metabolic Equivalent of Task (MET) in hours per week across all types of activity. Scores of over 168 hours per week were deemed implausible and coded as missing. See supplementary materials for more detail.

### Statistical analysis

Statistical analyses were conducted using R (version 4.2.2).

We compared sociodemographic and lifestyle variables for individuals with and without ACEs (*Table 1)* for the full sample and separately for females and males. In the full sample, diet, alcohol intake, smoking status and physical activity levels were corrected for gender by including gender as a covariate in regression models. Associations with age and socioeconomic status were measured using t-tests. Associations with ethnicity were evaluated using a chi-squared test. Binomial logistic regression analysis regressing ACE on alcohol intake, smoking status and physical activity levels were used. The association between ACEs and diet was evaluated using ordinal logistic regression.

**Table 1.**
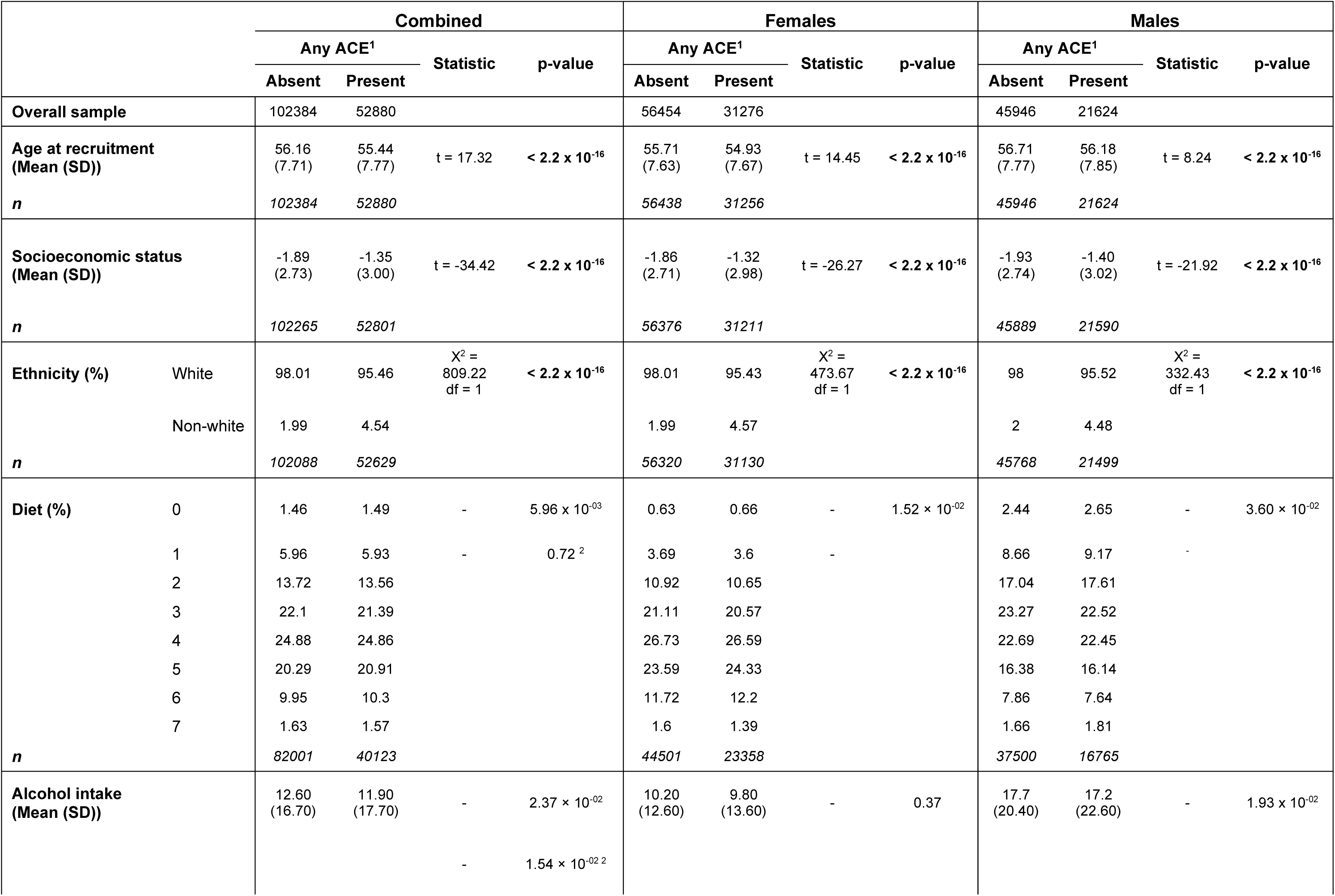
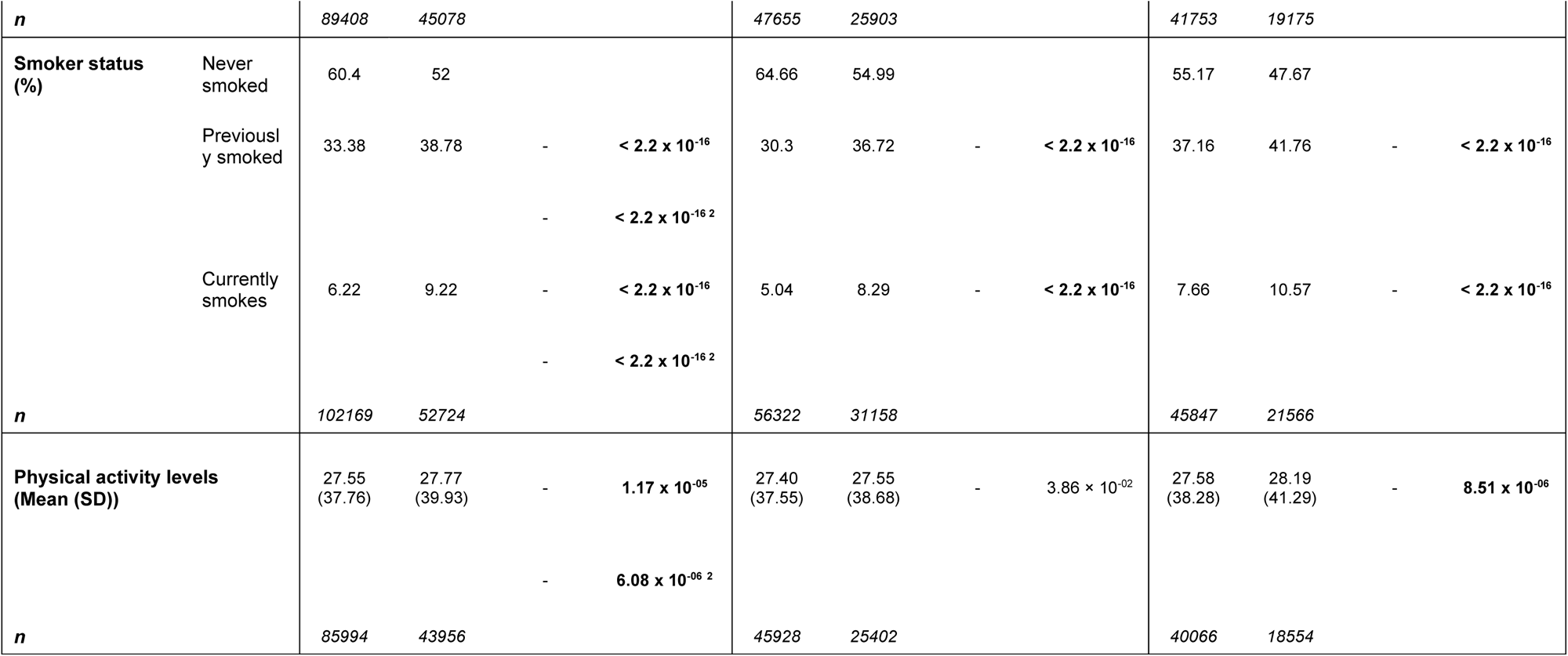
Prevalence of sociodemographic and lifestyle variables in individuals who do and do not have adverse childhood experiences (ACEs) in the full sample (lifestyle variables are corrected for gender) and separately by gender. All p-values are corrected for multiple testing using the Bonferroni Correction, raw p-values are reported and those formatted in bold survived correction for multiple testing. The adjusted alpha level for the tests using the combined sample (after accounting for eight tests) was 0.0063 and for males and females separately (after accounting for 13 tests) was 0.0038. ^1^Any ACE refers to reporting at least one of the following: emotional neglect, physical neglect, emotional abuse, physical abuse, sexual abuse. Associations with age and socioeconomic status were tested using parametric t-tests. Socioeconomic status is measured using Townsend Deprivation Index, where scores of 0 represent an area with overall mean values, positive values indicate areas with higher levels of material deprivation and negative values areas with lower levels of material deprivation. Association with ethnicity was evaluated using chi-squared test. Alcohol intake, smoker status and physical activity levels were tested using binomial logistic regression to ensure that significant differences between groups reflected changes in the mean as opposed to changes in the distribution. Alcohol intake is measured in units per week. Association with diet was tested using ordinal logistic regression. Higher diet scores indicate healthier diets. ^2^Regression models adjusted for gender. Gender coded as male = 0 and female = 1. SD = Standard Deviation.

To address aim 1, binomial logistic regressions were conducted regressing 13 individual and multimorbid health presentations on each type of ACE (*Table 2* and *Supplementary Tables 4-8*). Four binomial logistic regression models were tested: an unadjusted regression model (Model 1); followed by adjustment for demographic factors (age, gender, SES and ethnicity; Model 2), adjustment for lifestyle factors (diet, alcohol intake, smoking status and physical activity levels; Model 3) and adjustment for both demographic and lifestyle factors (Model 4). Models 2, 3 and 4 address Aim 2.

**Table 2.** Results of binary logistic regression models comparing the frequency of health outcomes in individuals with any ACE versus those without. ^1^Any ACE refers to reporting at least one of the following: emotional neglect, physical neglect, emotional abuse, physical abu se or sexual abuse. **^2^**Any IC refers to a diagnosis of depression and/or anxiety from primary or secondary electronic health care records (*Supplementary* Table 2). **^3^**Any CMC refers to a diagnosis of hypertension, T2D, obesity, CKD and/or dyslipidaemia from primary or secondary care records (*Supplementary* Table 2). **^4^**ICM-MM refers to a diagnosis of any IC plus any CMC from primary or secondary care records. CKD refers to chronic kidney disease (*Supplementary* Table 2). Model 1 is not adjusted for any covariates. Model 2 is adjusted for demographic variables: age, gender, ethnicity and socioeconomic status. Model 3 is adjusted for lifestyle variables: diet, alcohol intake, smoking status and physical activity levels. Model 4 is adjusted for demographic and lifestyle variables: age, gender, ethnicity, socioeconomic status, diet, alcohol intake, smoking status and physical activity levels. All p-values are corrected for multiple testing using the Bonferroni Correction, raw p-values are reported and those formatted in bold survived correction for multiple testing.

|  | Any ACE<sup>1</sup> |  |  |  |  |  |  |  |
| --- | --- | --- | --- | --- | --- | --- | --- | --- |
|  | Odds ratio<br>(95% CI) | p-value | Odds ratio<br>(95% CI) | p-value | Odds ratio<br>(95% CI) | p-value | Odds ratio<br>(95% CI) | p-value |
|  | Model 1<br>Unadjusted |  | Model 2<br>Adjusted for demographic<br>variables |  | Model 3<br>Adjusted for lifestyle variables |  | Model 4<br>Adjusted for demographic and<br>lifestyle variables |  |
| **Any IC<sup>2</sup>** | 1.84<br>(1.79-1.89) | **< 2.2 x 10<sup>-16</sup>** | 1.80<br>(1.75-1.85) | **< 2.2 x 10<sup>-16</sup>** | 1.70<br>(1.68-1.81) | **< 2.2 x 10<sup>-16</sup>** | 1.75<br>(1.68 - 1.82) | **< 2.2 x 10<sup>-16</sup>** |
| **Hypertension** | 1.08<br>(1.06-1.11) | **2.59 x 10<sup>-11</sup>** | 1.16<br>(1.13-1.19) | **< 2.2 x 10<sup>-16</sup>** | 1.07<br>(1.04-1.11) | **2.26 x 10<sup>-05</sup>** | 1.12<br>(1.08 - 1.16) | **2.13 x 10<sup>-11</sup>** |
| **T2D** | 1.29<br>(1.24-1.35) | **< 2.2 x 10<sup>-16</sup>** | 1.31<br>(1.25-1.37) | **< 2.2 x 10<sup>-16</sup>** | 1.24<br>(1.17-1.32) | **8.76 x 10<sup>-12</sup>** | 1.24<br>(1.16 - 1.32) | **2.42 x 10<sup>-11</sup>** |
| **Obesity** | 1.44<br>(1.38-1.49) | **< 2.2 x 10<sup>-16</sup>** | 1.41<br>(1.35-1.46) | **< 2.2 x 10<sup>-16</sup>** | 1.40<br>(1.32-1.47) | **< 2.2 x 10<sup>-16</sup>** | 1.38<br>(1.31 - 1.46) | **< 2.2 x 10<sup>-16</sup>** |
| **CKD** | 1.14<br>(1.08-1.20) | **1.07 x 10<sup>-06</sup>** | 1.20<br>(1.14-1.27) | **7.21 x 10<sup>-12</sup>** | 1.09<br>(1.01-1.17) | 1.95 x 10<sup>-02</sup> | 1.14<br>(1.06 - 1.23) | **4.05 x 10<sup>-04</sup>** |
| **Dyslipidaemia** | 1.11<br>(1.08-1.15) | **1.48 x 10<sup>-14</sup>** | 1.17<br>(1.14-1.21) | **< 2.2 x 10<sup>-16</sup>** | 1.09<br>(1.05-1.13) | **1.42 x 10<sup>-05</sup>** | 1.12<br>(1.08 - 1.16) | **1.1 x 10<sup>-08</sup>** |
| **Any CMC<sup>3</sup>** | 1.14<br>(1.11-1.16) | **< 2.2 x 10<sup>-16</sup>** | 1.21<br>(1.19-1.24) | **< 2.2 x 10<sup>-16</sup>** | 1.13<br>(1.09-1.16) | **1.29 x 10<sup>-15</sup>** | 1.18<br>(1.14 - 1.22) | **< 2.2 x 10<sup>-16</sup>** |
| **Any IC plus hypertension** | 1.73<br>(1.66-1.81) | **< 2.2 x 10<sup>-16</sup>** | 1.75<br>(1.67-1.83) | **< 2.2 x 10<sup>-16</sup>** | 1.64<br>(1.54-1.75) | **< 2.2 x 10<sup>-16</sup>** | 1.67<br>(1.57 - 1.78) | **< 2.2 x 10<sup>-16</sup>** |
| **Any IC plus T2D** | 1.97<br>(1.80-2.16) | **< 2.2 x 10<sup>-16</sup>** | 1.90<br>(1.74-2.09) | **< 2.2 x 10<sup>-16</sup>** | 1.82<br>(1.60-2.07) | **< 2.2 x 10<sup>-16</sup>** | 1.79<br>(1.57 - 2.04) | **< 2.2 x 10<sup>-16</sup>** |
| **Any IC plus obesity** | 2.15<br>(2.01-2.29) | **< 2.2 x 10<sup>-16</sup>** | 2.01<br>(1.88-2.15) | **< 2.2 x 10<sup>-16</sup>** | 1.99<br>(1.80-2.19) | **< 2.2 x 10<sup>-16</sup>** | 1.93<br>(1.75 - 2.13) | **< 2.2 x 10<sup>-16</sup>** |
| **Any IC plus CKD** | 1.86<br>(1.68-2.05) | **< 2.2 x 10<sup>-16</sup>** | 1.90<br>(1.72-2.10) | **< 2.2 x 10<sup>-16</sup>** | 1.62<br>(1.40-1.87) | **3.60 x 10<sup>-11</sup>** | 1.67<br>(1.45 - 1.93) | **2.75 x 10<sup>-12</sup>** |
| **Any IC plus dyslipidaemia** | 1.78<br>(1.69-1.87) | **< 2.2 x 10<sup>-16</sup>** | 1.78<br>(1.69-1.88) | **< 2.2 x 10<sup>-16</sup>** | 1.67<br>(1.55-1.79) | **< 2.2 x 10<sup>-16</sup>** | 1.69<br>(1.57 - 1.82) | **< 2.2 x 10<sup>-16</sup>** |
| **Any ICM-MM<sup>4</sup>** | 1.81<br>(1.74-1.88) | **< 2.2 x 10<sup>-16</sup>** | 1.79<br>(1.73-1.86) | **< 2.2 x 10<sup>-16</sup>** | 1.72<br>(1.63-1.81) | **< 2.2 x 10<sup>-16</sup>** | 1.73<br>(1.64 - 1.83) | **< 2.2 x 10<sup>-16</sup>** |

To examine whether the impact of the ACEs on health conditions differed by gender (aim 3), binomial logistic regressions were conducted regressing health conditions on each type of ACE in females and males separately. The significance of the difference in odds ratios (ORs) between genders was tested by adding an interaction term (gender * ACE) to the regression analyses in the combined sample (*Table 3* and *Supplementary Tables 9-13*).

**Table 3.** Results of binary logistic regression models comparing the frequency of health outcomes in those with any ACE and those without in females and males separately. ^1^Any ACE refers to reporting at least one of the following: emotional neglect, physical neglect, emotional abuse, physical abuse or sexual abuse. ^2^Any IC refers to a diagnosis of depression and/or anxiety from primary or secondary care records (*Supplementary* Table 2). ^3^Any CMC refers to a diagnosis of hypertension, T2D, obesity, CKD and/or dyslipidaemia from primary or secondary care records (*Supplementary* Table 2). ^4^ICM-MM refers to a diagnosis of any IC plus any CMC from primary or secondary care records. CKD refers to chronic kidney disease (*Supplementary* Table 2). ^5,6^Odds ratios and confidence intervals, adjusted for age, ethnicity, socioeconomic status, diet, alcohol intake, smoking status and physical activity levels, are reported for females and males separately. ^7^P-values of the interaction between any ACE and gender, adjusted for age, ethnicity, socioeconomic status, diet, alcohol intake, smoking status and physical activity levels, are reported. All p-values are corrected for multiple testing using the Bonferroni Correction, raw p-values are reported and none survived correction for multiple testing.

| Health outcomes | Any ACE<sup>1</sup> |  |  |
| --- | --- | --- | --- |
|  | Odds ratio<sup>5</sup><br>(Females)<br>(95% CI) | Odds ratio<sup>6</sup><br>(Males)<br>(95% CI) | p-value of<br>difference<sup>7</sup> |
| Any IC<sup>2</sup> | 1.73<br>(1.64-1.82) | 1.71<br>(1.60-1.82) | 0.94 |
| Hypertension | 1.16<br>(1.11-1.22) | 1.12<br>(1.07-1.18) | 0.74 |
| T2D | 1.34<br>(1.20-1.49) | 1.25<br>(1.15-1.35) | 0.40 |
| Obesity | 1.45<br>(1.34-1.56) | 1.31<br>(1.21-1.42) | 0.07 |
| CKD | 1.17<br>(1.05-1.30) | 1.13<br>(1.02-1.24) | 0.80 |
| Dyslipidaemia | 1.19<br>(1.12-1.26) | 1.11<br>(1.06-1.17) | 0.17 |
| Any CMC<sup>3</sup> | 1.24<br>(1.18-1.29) | 1.16<br>(1.11-1.22) | 0.13 |
| Any IC plus hypertension | 1.70<br>(1.56-1.85) | 1.62<br>(1.48-1.78) | 0.68 |
| Any IC plus T2D | 1.72<br>(1.41-2.09) | 1.88<br>(1.57-2.24) | 0.59 |
| Any IC plus obesity | 1.89<br>(1.67-2.13) | 1.91<br>(1.62-2.26) | 0.94 |
| Any IC plus CKD | 1.75<br>(1.45-2.10) | 1.52<br>(1.21-1.91) | 0.51 |
| Any IC plus dyslipidaemia | 1.67<br>(1.51-1.85) | 1.69<br>(1.52-1.88) | 0.77 |
| Any ICM-MM<sup>4</sup> | 1.74<br>(1.62-1.86) | 1.68<br>(1.55-1.82) | 0.70 |

For *Tables 2 and 3* (and *Supplementary Tables 4-13*) p-values are corrected for multiple testing using the Bonferroni correction, the adjusted alpha level after accounting for six ACE categories and 13 health conditions was 0.00064.

## Results

Rates of ACEs in the total sample, and for females and males separately, are presented in *Supplementary Table 3.* 33.64% of the sample reported at least one ACE, and 5.41% three or more ACEs. The prevalences of emotional and physical neglect, and emotional and sexual abuse were significantly higher in females than males (all p-values < 2.2 x 10^-16^), but there was no difference for physical abuse. Emotional neglect was the most prevalent ACE for both females (23.15%) and males (21.67%), whilst physical neglect was the least prevalent ACE for both females (6.15%) and males (5.25%).

*Table 1* presents the sociodemographic and lifestyle characteristics of those who reported ACEs compared to those who did not. The findings are presented separately for females and males as well as combined (taking gender into account as a covariate for the lifestyle variables). Those with any ACE were younger than those without by approximately 8 months (t(106084) = 17.32, *p* < 2.2 x 10^-16^). Presence of any ACE was associated with lower socio-economic status (t(98187) = -34.42, *p* < 2.2 x 10^-16^)). There was a higher prevalence of people of non-white ethnic backgrounds in individuals with any ACE (4.54%) than without (1.99%) (χ² (1, N = 156629) = 809.22, *p* < 2.2 x 10^-16^). Compared to those who had never smoked, there was a higher risk of experiencing any ACE when the participant was a past or current smoker (past smoker OR = 1.37, *p* < 2.2 x 10^-16^; current smoker OR = 1.77, *p* < 2.2 x 10^-16^). Finally, those with any ACE had increased mean physical activity levels compared to those without ACEs, however the effect size and absolute difference in mean was minimal (combined sample OR = 1.001, *p* = 6.08 x 10^-06^). All the associations were present in both females and males, except for physical activity, which was only present in males (OR = 1.001, *p* = 8.51 x 10^-06^).

We next examined whether reporting any ACE was associated with risk of health presentations (Aim 1) (*Table 2*). We found associations with all individual and multimorbid presentations in Model 1 (unadjusted model). The associations were stronger between ACEs and any IC and any ICM-MM respectively, versus ACEs and any CMC. For example, the OR of being diagnosed with any IC after experiencing any ACE was 1.84, (*p* < 2.2 x 10^-16^), whereas for individual CMC conditions the ORs ranged from 1.08-1.44, with obesity showing the strongest association (OR = 1.44, *p* < 2.2 x 10^-16^). For any CMC, the OR was 1.14, (*p* = < 2.2 x 10^-16^). For pairwise ICM-MM combinations, ORs ranged from 1.73 (*p* < 2.2 x 10^-16^) for IC plus hypertension to 2.15 (*p* < 2.2 x 10^-16^) for IC plus obesity. Association with any ICM-MM (OR= 1.81, *p* < 2.2 x 10^-16^) was similar to that of any ACE and any IC, and broadly in line with multimorbid presentations.

We subsequently adjusted for demographic (Model 2) and lifestyle factors (Model 3) as well as both (Model 4) (aim 2). Similar associations to Model 1 were found in Models 2 and 4, whereas in Model 3, one difference was found: any ACE was no longer associated with CKD. A stepwise regression model indicated this reduction in odds ratio and loss of significance was attributable to the inclusion of smoking status (*p* = 2.66 x 10^-3^ : not statistically significant after multiple correction tests).

Unadjusted associations of individual ACEs and health conditions are reported in *Supplementary Tables 4-8,* Model 1 (aim 1). Physical neglect and physical abuse were associated with all individual and multimorbid presentations. Emotional neglect was associated with all conditions except for CKD, emotional abuse was associated with all conditions except for hypertension and CKD and sexual abuse was only associated with any IC, T2D, obesity and all multimorbid presentations. *Supplementary Tables 4-8* also provide results for Model 2, 3 and 4 (Aim 2), and present that the associations remain largely unaffected by the inclusion of demographic and lifestyle factors. All individual ACEs were most strongly associated with IC plus obesity, except for sexual abuse which was most strongly associated with IC plus CKD (Model 4, OR = 1.73, *p* = 1.71 x 10^-07^). Generally, age and gender were key drivers of changes in statistical significance across models, whereby older and male participants were at higher risk of individual and multimorbid presentations.

*Table 3* presents associations between ACEs and health conditions for females and males separately (aim 3). Presence of any ACE was associated with increased risk of all health presentations in both females and males. There was no evidence that the association between any ACE and any individual or multimorbid presentations varied by gender.

Results for all individual ACE types and health presentations (*Supplementary Tables 9-13*) indicated no gender differences, except for emotional neglect. Although both females and males have increased risk of any CMC after reporting emotional neglect, females have higher odds of any CMC diagnosis than males (female OR = 1.28, male OR = 1.11, *p* = 1.25 x 10^-04^).

## Discussion

This is the first study to examine the impact of childhood adversity on the multimorbid presentation of IC and a range of CMCs, which confer risk for later cardiovascular disease. We studied the associations between a range of ACEs and diagnoses of any IC, T2D, obesity, hypertension, dyslipidaemia, CKD, any CMC and their multimorbid combinations in a large population-based cohort of mid to older age people. We found that experiences of a range of different ACEs increased the risk of all individual and multimorbid presentations, with similar findings for females and males. Furthermore, the findings generally remained the same after adjustment for demographic and lifestyle variables. These findings highlight the impact of ACEs on complex and difficult to treat chronic health presentations in older age and have implications for prevention and early intervention efforts.

The presence of any ACE increased the risk of any IC by 75%, any CMC by 18% and any ICM-MM by 73% even when accounting for demographic and lifestyle factors. This work builds on prior studies by extending the investigation to an older population and shifting the focus from counts of long-term conditions or disease categories to specific combinations of ICs and CMCs. Importantly, the current study used formal diagnoses recorded in EHRs. These results align, however, with previous research reporting that ACEs are associated with increased likelihood of IC (Merrick et al., 2017), CMC (Suglia et al., 2018) and mental and physical health multimorbidity including ICs and CMCs (Hanlon et al., 2020; Taylor & Demakakos, 2024). The results suggest that ACEs have a stronger relationship with IC and multimorbid presentations than individual CMCs, confirming and extending findings by Hughes et al. (2017). The same is true across all individual ACE types.

The presence of any ACE was most strongly associated with the combination of IC and obesity which is consistent with previous studies finding IC and obesity as comorbid outcomes of ACEs (Davies et al., 2024; Taylor & Demakakos, 2024). Obesity is a trait with complex multifactorial aetiologies, therefore associations between IC and obesity are likely to be complicated and nuanced (Masood & Moorthy, 2023). Possible theories range from the notion that ACEs can motivate people to gain weight to change their body from the one they were abused in (Felitti et al., 1998) and a probable casual bidirectional relationship between BMI and depression (Badillo, Khatib, Kahar, & Khanna, 2022; Steptoe & Frank, 2023).

The patterns of associations tended to be similar across the five different ACEs. All individual ACEs were significantly associated with any IC and all multimorbid combinations of IC and CMC. IC plus obesity, IC plus T2D and IC plus CKD were most strongly associated with individual ACEs, suggesting those who have experienced ACEs might be particularly vulnerable to these conditions as they age. One theory for this association is that chronic stress, which often accompanies adverse childhood experiences, can lead to allostatic load and constant arousal of hypothalamic-pituitary-adrenal (HPA) axis (Kalmakis, Meyer, Chiodo, & Leung, 2015). This in turn may disrupt the cortisol levels in the body, which can be a biomarker of anxiety and depression (Brindle, Pearson, & Ginty, 2022). Disruption of cortisol regulation can also increase food intake and cravings for sugar and salt, giving rise to excessive consumption as a coping mechanism with consequences for physical health (Epel, Lapidus, McEwen, & Brownell, 2001; Jackson, Kirschbaum, & Steptoe, 2017; Testa et al., 2024). Furthermore, both anxiety and depression have been directly related to overeating and binge eating as a form of coping with poor mental health (Torres & Nowson, 2007).

Reporting any ACE increased the likelihood of a diagnosis of IC, CMC and ICM-MM in both females and males. One finding indicated a sex difference: females who had experienced emotional neglect had greater risk of any CMC than males. However, overall, this study highlights that both genders have similar risks of poor health in mid to late adulthood after experiencing ACEs, including ICs. This novel finding adds to a literature that has predominantly focused on females or combined samples.

Those with an ACE were, on average, younger, living in more economically deprived areas and less likely to be of European ancestry, highlighting that the experience of ACEs is socially and culturally patterned as confirmed by previous studies (Merrick, 2019; Metzler, Merrick, Klevens, Ports, & Ford, 2017). Research in populations with a more diverse range of ethnic backgrounds is needed to further establish the relationship between ACEs and ethnicity.

Research has indicated that habitual lifestyle health risk behaviours such as smoking, alcohol use, poor diet and sedentary lifestyle could account for the enduring impact of ACEs on physical and mental health (Bellis, Lowey, Leckenby, Hughes, & Harrison, 2014). In this study, people with ACEs were more likely to be current or past smokers, confirming that ACEs may increase engagement in health risk behaviours. We also found associations between higher physical activity levels and ACEs; however, the effect sizes were small and including physical activity levels as a covariate in analysis had little impact on the findings. Alcohol intake was not associated with ACEs, potentially indicative of the age of UK Biobank participants. Levels of alcohol intake decrease in later adulthood (Britton, Ben-Shlomo, Benzeval, Kuh, & Bell, 2015) and it may be possible that stronger associations would be present in younger cohorts (Zhen-Duan, Colombo, Cruz-Gonzalez, Hoyos, & Alvarez, 2023). We did not find evidence that diet was associated with ACEs.

### Strengths and Limitations

Important strengths of this study include the large sample size, use of linked EHR data allowing for formal diagnoses of health conditions and the availability of key covariates such as lifestyle variables. However, there are limitations that must be acknowledged. It is possible there may be reporting biases such that individuals who are most affected by their trauma may have responded “Prefer not to say” to the CTS. We coded these responses as missing and thus these findings may represent an underestimate of the impact of ACEs on mental and physical health in older age.

Compared to the general population, UK Biobank participants are more affluent (Fry et al., 2017). In this study, although those with ACEs were more deprived than those without, the whole sample was still less materially deprived relative to the UK national average. UK Biobank participants are also less likely to have ethnic minority status, or to be obese, smoke, drink alcohol daily and have fewer self-reported health conditions (Fry et al., 2017). Those that continued to participate in the follow-up MHQ (which included the ACEs) were better educated, healthier, of a higher socioeconomic status and had lower rates of smoking than those who did not (Davis et al., 2020). Importantly, despite these biases, associations between ACEs and health conditions remained, suggesting that in the general population, where there may be increased vulnerability to ACEs and poor later health, these associations may be stronger.

The use of EHRs ensured the results were not biased by self-report, such as social desirability bias or recall bias. However, EHRs were designed to support clinical practice rather than research and therefore have limitations such as selective recording of conditions which may produce incomplete or misrepresentative data. Mental health diagnoses, particularly, are poorly captured in EHRs (Madden, Lakoma, Rusinak, Lu, & Soumerai, 2016). Furthermore, UK Biobank has incomplete coverage of primary care records and many of the conditions examined are managed in primary care, suggesting that the recorded frequencies of diagnoses may be lower than the true frequencies.

The ACEs studied in this work are based on retrospective reports and may be subject to recall bias such that those experiencing trauma may find it difficult to access autobiographical memories (Moore & Zoellner, 2007) and that those who are currently euthymic may find it difficult to recall negative memories (Colman et al., 2016). Furthermore, there are differences in associations of ACEs and psychopathology depending on whether the ACE is assessed retrospectively or prospectively, which highlights that subjective memory is potentially important in explaining associations between ACEs and IC in this study (Danese & Widom, 2020).

Finally, the fact that stronger associations were found with IC and multimorbid presentations than individual CMCs raises the question of whether there are direct pathways from ACEs to IC and CMC respectively, or whether ACEs increase the risk of ICs which in turn increase the risk of CMCs. Investigation into this was beyond the scope of the study because it would require conditioning on those who had any IC thereby creating risk of collider bias and spurious associations (Holmberg & Andersen, 2022).

### Clinical implications

The results of this study amplify recent claims that ACEs should be treated as a public health issue (Hughes et al., 2017). Those who have experienced ACEs may prefer to be aware of the risk of developing IC, CMC and ICM-MM in older age, allowing for self-advocacy in clinical settings and motivating the adoption of healthy lifestyle habits throughout life to support later wellbeing. Importantly, risk prediction within the patient and public domain should be handled with care, due to the risk of stigmatisation and self-fulfilling prophecy (Stephens et al., 2023). Males may experience additional barriers to service-seeking after trauma such as fear of disclosure and challenge to masculinity (Denhard et al., 2024). Therefore, an emphasis should be placed on motivating males to actively seek similar support after trauma as this study highlights that both females and males have similar risk of experiencing IC after ACEs. This research presents evidence that ACEs contribute to multimorbidity, a major driver of costs incurred by the public health services worldwide. At the population level, faster detection and intervention of ACEs by public health services may reduce cases of multimorbidity and allow for a decrease in healthcare costs. Moreover, joining up public health services, creating one integrated system of care, could allow for more effective information sharing, giving health care providers context for better decision making and individualised care.

The implications of the presence of ACEs for clinical practice remain unclear. The literature on the implementation of routine ACE enquiry in primary care has highlighted a number of caveats such as an over-reliance on crude ACE scores, which do not account for the subjective experience of different ACE types, and a lack of training on safely implementing the enquiry in primary care (Anda, Porter, & Brown, 2020; Gentry & Paterson, 2022; Hardcastle & Bellis, 2018; Loveday et al., 2022; McLennan, Gonzalez, MacMillan, & Afifi, 2024). A focus on strengthening patient-clinician interactions through connection, longer appointment times in which trust can be built and training on safely discussing trauma, may be more beneficial (Jones, Merrick, & Houry, 2020; Zulman et al., 2020).

Finally, it should be acknowledged that a substantial number of people go on to healthy and fulfilling adulthoods after having experienced ACEs, and their lived experience could be drawn upon to develop interventions that improve later health outcomes. For example, there is a growing body of literature on the concept of “continuing adversity” (those who experience prolonged adversity after childhood have worse outcomes compared to those whose trauma is confined to childhood (Horwitz, Widom, McLaughlin, & White, 2001)), and the importance of improving resilience after experiencing ACEs to mediate poor health outcomes in childhood and adulthood (Haczkewicz et al., 2024).

## Conclusion

This study shows that ACEs impact IC, CMC and ICM-MM in older adulthood and demographic and lifestyle factors do not change these associations. Males are at similar risk as females of poor health in adulthood following ACEs. The importance of early detection and intervention to prevent ACEs and the use of trauma-informed care cannot be understated.

## Data Availability

All data produced are available online at https://www.ukbiobank.ac.uk/use-our-data/

## Acknowledgements

This research has been conducted using the UK Biobank Resource under application number 79704. Full list of LINC members: Marianne B. M. van den Bree, George Kirov, Michael J. Owen, James T. R. Walters, Peter A. Holmans, Jane Lynch, Ioanna K. Katzourou, Lowri O’Donovan (Cardiff University, UK). David A. van Heel, Sarah Finer, Daniel Stow (Queen Mary University of London, UK). Golam M. Khandaker, Nicholas J. Timpson, John A. A. MacLeod, Julie P. Clayton, Ruby S. M. Tsang, Jane Sprackman, Shahid Khan (University of Bristol, UK). Inês Barroso, Rupert A. Payne (University of Exeter, UK). Mark Mon-Williams, Megan L. Wood (University of Leeds, UK). Hilary C. Martin (Wellcome Sanger Institute, UK). Thomas Werge, Andrés Ingason (Institute of Biological Psychiatry, Denmark).

We would like to acknowledge the LIfespaN multimorbidity research Collaborative (LINC) patient and public involvement group for their input on this research.

## Financial contributions

This work was funded by a PhD studentship to LKB from Health and Care Research Wales (MvdB; PH, MJO, FR, MMW; HS 22 04) and the Tackling Multimorbidity at Scale Strategic Priorities Fund programme (MvdB, PH, MJO, MMW, RP; MR/W014416/1) delivered by the Medical Research Council and the National Institute for Health Research in partnership with the Economic and Social Research Council and in collaboration with the Engineering and Physical Sciences Research Council.

## Ethical standards

The authors assert that all procedures contributing to this work comply with the ethical standards of the relevant national and institutional committees on human experimentation and with the Helsinki Declaration of 1975, as revised in 2008.

## Competing interests

Michael J. Owen reports grants from Akrivia Health and Takeda Pharmaceuticals outside the submitted work.

## References

Academy of Medical Sciences. (2018). Multimorbidity: a priority for global health research. Retrieved from Anda, R. F., Porter, L. E., & Brown, D. W. (2020). Inside the adverse childhood experience score: Strengths, limitations, and misapplications. *American journal of preventive medicine*, 59(2), 293–295. Retrieved from https://www.sciencedirect.com/science/article/pii/S0749379720300581?via%3Dihub

Badillo, N., Khatib, M., Kahar, P., & Khanna, D. (2022). Correlation between body mass index and depression/depression-like symptoms among different genders and races. Cureus, 14(2).

Beesdo, K., Knappe, S., & Pine, D. S. (2009). Anxiety and anxiety disorders in children and adolescents: developmental issues and implications for DSM-V. The Psychiatric Clinics of North America, 32(3), 483. Retrieved from https://pmc.ncbi.nlm.nih.gov/articles/PMC3018839/

Bellis, M. A., Lowey, H., Leckenby, N., Hughes, K., & Harrison, D. (2014). Adverse childhood experiences: retrospective study to determine their impact on adult health behaviours and health outcomes in a UK population. Journal of public health, 36(1), 81–91. Retrieved from https://watermark.silverchair.com/fdt038.pdf?token=AQECAHi208BE49Ooan9kkhW_Ercy7Dm3ZL_9Cf3qfKAc485ysgAAA2UwggNhBgkqhkiG9w0BBwagggNSMIIDTgIBADCCA0cGCSqGSIb3DQEHATAeBglghkgBZQMEAS4wEQQM-EQ8sd-DbAdOQSb8AgEQgIIDGBPCKyOC_QDub2W0YqZ-QRSKra95W9I4kEoDGRTdyMhtVVYpGcGIK478EL64CFvwqhQpFtl2W3vUBnelQA-9MerRkteBhPKPoVEJ5PZzWVstK5Dk8hBIMBA7XCEmQpR2weE7nO_cJAquRNkDxPKhSHAqjHtPS1TEY_Vzk0aTJtY3S5ifLuAsp5Sf1mWMyB6cMXCywX5J5F3OjhoN3PFQWVXnBfGktIWxmMJuIzRNxSLKNPgQdK4K6I-TBUign8Q1rD3Cubl0H1yAqdwiyiAvr81t_tdxDoXUwgZkjjnDmTHwpbjGzliYhC9un1wiDlMGLs3RXvp7-01ptnaDwaEbeAkCbQVxXCeGrjg4m1SOCqki34u1JBCRQ4Nbx_DjbVMqkgSiEt10kbTuG8iNPK6Mm9FxMFkoKYBCDwLy5_AAsOVS2bnceqSJJ2fOmMFCFvmMbCP-9qhkcoNitc-nKxuqy_se2oRIoglQb45IBMNU3-fWIX62DvAAmFnc-YaU-5mIM2ghwWwMmBQkb8ennRbj9r7MQUb_DHKRmOWayEYUCRdDgUv_NCTsG8ku3Jh5xhBS63HiQZE3la9ZLomT4kLOr7uh5Km9VVr6jEe23SZqy6bDEvXKUsjOQXA_KDJ2SwzsYI-JfBBz3JRTAYxivm6-uKX8ogpUUExsUWrwTsWSyXLhBYazaHOfuHpQIbqy3243mhuc3dabPbS_0lRz2RmlImhsC94HZMMK97JJbbnX-Bn2gUkzRfLBRH4YRAj5EFgjPR_sSwIDLDn8qTOVDNENRiIOewwoJTzgTxSmjEDu52swWQbcHm59gadNySmmVERHBhSgl2ydI-mKVrs8YncdImxAarAcIL-79f3XtMUxR4E7ZUKHxmR_k83DpnJ4zTNmb-GWbFRIvXxOdbSdhzd_IMiVi6Ju7vldTrG-I9vQCpftZOuaqChOiELHSR7gzzMEqTJCU8fc6b4WiQa8_aA7DfKIIX-WaZc8YpTTWiiwR97vvJFs9T3d1tgt-Mc6tx85_8SxVlHEg-9VNmjABYLUp8CYpQg44oatBg

Berenson, G. S., Srinivasan, S. R., Hunter, S. M., Nicklas, T. A., Freedman, D. S., Shear, C. L., & Webber, L. S. (1989). Risk factors in early life as predictors of adult heart disease: the Bogalusa Heart Study. The American journal of the medical sciences, 298(3), 141–151. Retrieved from https://www.sciencedirect.com/science/article/abs/pii/S0002962915362418?via%3Dihub

Blissett, J. (2011). Relationships between parenting style, feeding style and feeding practices and fruit and vegetable consumption in early childhood. Appetite, 57(3), 826–831. Retrieved from https://www.sciencedirect.com/science/article/abs/pii/S0195666311004752?via%3Dihub https://www.sciencedirect.com/science/article/pii/S0195666311004752?via%3Dihub

Brindle, R. C., Pearson, A., & Ginty, A. T. (2022). Adverse childhood experiences (ACEs) relate to blunted cardiovascular and cortisol reactivity to acute laboratory stress: A systematic review and meta-analysis. Neuroscience & Biobehavioral Reviews, 134, 104530.

Britton, A., Ben-Shlomo, Y., Benzeval, M., Kuh, D., & Bell, S. (2015). Life course trajectories of alcohol consumption in the United Kingdom using longitudinal data from nine cohort studies. BMC medicine, 13(1), 47.

Bycroft, C., Freeman, C., Petkova, D., Band, G., Elliott, L. T., Sharp, K., . . . O’Connell, J. (2018). The UK Biobank resource with deep phenotyping and genomic data. Nature, 562(7726), 203–209. Retrieved from https://www.nature.com/articles/s41586-018-0579-z.pdf

Carlson, D. M., & Yarns, B. C. (2023). Managing medical and psychiatric multimorbidity in older patients. Therapeutic Advances in Psychopharmacology, 13, 20451253231195274. Retrieved from https://pmc.ncbi.nlm.nih.gov/articles/PMC10469275/pdf/10.1177_20451253231195274.pdf

Chowdhury, S. R., Das, D. C., Sunna, T. C., Beyene, J., & Hossain, A. (2023). Global and regional prevalence of multimorbidity in the adult population in community settings: a systematic review and meta-analysis. EClinicalMedicine, 57.

Colman, I., Kingsbury, M., Garad, Y., Zeng, Y., Naicker, K., Patten, S., . . . Thompson, A. H. (2016). Consistency in adult reporting of adverse childhood experiences. Psychological medicine, 46(3), 543–549. Retrieved from https://www.cambridge.org/core/services/aop-cambridge-core/content/view/0F94F0451B1CCB80699660E066258E6C/S0033291715002032a.pdf/div-class-title-consistency-in-adult-reporting-of-adverse-childhood-experiences-div.pdf

Danese, A., & Widom, C. S. (2020). Objective and subjective experiences of child maltreatment and their relationships with psychopathology. Nature human behaviour, 4(8), 811–818. Retrieved from https://www.nature.com/articles/s41562-020-0880-3.pdf

Davies, J. L., Lawrence, D., Bagshaw, R., Watt, A., Mills, S., & Seage, C. H. (2024). Psychological trauma predicts obesity in Welsh Secure mental health inpatients. International Journal of Forensic Mental Health, 23(3), 241–250.

Davis, K. A., Coleman, J. R., Adams, M., Allen, N., Breen, G., Cullen, B., . . . Holliday, J. (2020). Mental health in UK Biobank–development, implementation and results from an online questionnaire completed by 157 366 participants: a reanalysis. BJPsych Open, 6(2), e18. Retrieved from https://www.cambridge.org/core/services/aop-cambridge-core/content/view/F402F460E7731030354A07F9AD8F46A1/S2056472419001005a.pdf/div-class-title-mental-health-in-uk-biobank-development-implementation-and-results-from-an-online-questionnaire-completed-by-157-366-participants-a-reanalysis-div.pdf

Denhard, L., Brown, C., Kanagasabai, U., Thorsen, V., Kambona, C., Kamagate, F., . . . McOwen, J. (2024). Service-seeking behaviors among male victims of violence in five African countries: The effects of positive and adverse childhood experiences. Child Abuse & Neglect, 150, 106452.

Epel, E., Lapidus, R., McEwen, B., & Brownell, K. (2001). Stress may add bite to appetite in women: a laboratory study of stress-induced cortisol and eating behavior. Psychoneuroendocrinology, 26(1), 37–49. Retrieved from https://www.sciencedirect.com/science/article/abs/pii/S0306453000000354?via%3Dihub https://www.sciencedirect.com/science/article/pii/S0306453000000354?via%3Dihub

Felitti, V. J., Anda, R. F., Nordenberg, D., Williamson, D. F., Spitz, A. M., Edwards, V., & Marks, J. S. (1998). Relationship of childhood abuse and household dysfunction to many of the leading causes of death in adults: The Adverse Childhood Experiences (ACE) Study. American journal of preventive medicine, 14(4), 245–258. Retrieved from https://www.ajpmonline.org/article/S0749-3797(98)00017-8/pdf

Fry, A., Littlejohns, T. J., Sudlow, C., Doherty, N., Adamska, L., Sprosen, T., . . . Allen, N. E. (2017). Comparison of sociodemographic and health-related characteristics of UK Biobank participants with those of the general population. American journal of epidemiology, 186(9), 1026–1034. Retrieved from https://pmc.ncbi.nlm.nih.gov/articles/PMC5860371/pdf/kwx246.pdf

Garber, J., Brunwasser, S. M., Zerr, A. A., Schwartz, K. T., Sova, K., & Weersing, V. R. (2016). Treatment and prevention of depression and anxiety in youth: test of cross-over effects. Depression and anxiety, 33(10), 939–959. Retrieved from https://onlinelibrary.wiley.com/doi/pdfdirect/10.1002/da.22519?download=true

Gentry, S., & Paterson, B. (2022). Does screening or routine enquiry for adverse childhood experiences (ACEs) meet criteria for a screening programme? A rapid evidence summary. Journal of public health, 44(4), 810–822. Retrieved from https://watermark.silverchair.com/fdab238.pdf?token=AQECAHi208BE49Ooan9kkhW_Ercy7Dm3ZL_9Cf3qfKAc485ysgAAA2gwggNkBgkqhkiG9w0BBwagggNVMIIDUQIBADCCA0oGCSqGSIb3DQEHATAeBglghkgBZQMEAS4wEQQMxLj5nzjQWZJElZuEAgEQgIIDG8N-f18qW_BU3WP4yGeI7Y_a8Otj_8nB40f3WBCw6dLpR6vo32TnzFdVLIgZK4Ss-Ht0aUTQD_ePG-7De7PfcfNjFHmHGBRqWi3XowSjncjwB2mGLV4B7gnqnURxXLV9rO-LD4sc6BpX0K3N5h_HjKIDaC-xQu2kdRlaDreBUICXlwY1fePxP3XLWhOk-gi9mcWDj5rJzxGW_0R5g9BDEillLkQ5LyW2rljO4LhF9IpPiGZ5UnLvM6E7IlROMF7i0W9X1L--ixna262wV8egmnFQL_Q_oeSXlLQgpUqrcyq8dB-BeRWXEUCqke7z2xjEuT1gjJeUoY4eroZdHsWRFIRCpzzt3dtBoxeAzw7BIw999vtne35Juo8KrAdhvA62uW_SiUsJxvfYs4IcK463-lez3_H1B09N2AB7jIeDDaJhBKzSPOz2w8oclRSHk18z-wbC34IJqGnAdFGYQgmHHJ_9voArcQyZp6qYNirDtedtuwo6zOBdog_w8UDUbujaU7eFsLBCxfouKUGUz_Sof8mLa1sZyZ4NnrPkherVnnGM-Ac-Ytx1IPknECWpuq0fmgM4JvF2WFF2bSOG411LdKwq2OYMKvP-JphMcrWS1b74m31RAY9sqjAygLRogKZa8viXlsutlUIZWinfeU2Id4fdhQAdrH8FuvfgfiJhXZ9IbF970gjAWiGFeyZzl2T5ed0ENWO-lIteZMhEuw_0q-fDj5kuStbI8ZWjnWbf79Xju1J2VV0coHtPv1O6lxVZPlyEg3kS0BSD2qSjLnxrkCCPlqzRcSe-G-iIT20-io02VFN8zrjarqF3n5suaV5EmScvF356qEl0owk2uZkBMERnlUOM_QvNVaHDk2jugDaYOk9QPa0g_bKjwLMtLLlFPMaMqGwbEy-xiensFxpIxKSa3QL_hHqNwBluaLSPJzWUqfcdJh7c32jkmhjKguWy_a9bJ9XTLJe_WpYmMBdb5rK4Ar5JnPXd1h8ufE_4kBxkEOzpuXVbowC8XHLU5N5zBtuzFFZdxAB08EVYWlbrHYIx3tytVxlUTWFRUw

Goodwin, G. M., & Stein, D. J. (2021). Generalised anxiety disorder and depression: contemporary treatment approaches. Advances in therapy, 38(Suppl 2), 45–51. Retrieved from https://link.springer.com/content/pdf/10.1007/s12325-021-01859-8.pdf

Grabe, H. J., Schulz, A., Schmidt, C. O., Appel, K., Driessen, M., Wingenfeld, K., . . . Berger, K. (2012). A brief instrument for the assessment of childhood abuse and neglect: the childhood trauma screener (CTS). Psychiatrische Praxis, 39(3), 109–115.

Haahr-Pedersen, I., Perera, C., Hyland, P., Vallières, F., Murphy, D., Hansen, M., . . . Cloitre, M. (2020). Females have more complex patterns of childhood adversity: Implications for mental, social, and emotional outcomes in adulthood. European Journal of Psychotraumatology, 11(1), 1708618. Retrieved from https://pmc.ncbi.nlm.nih.gov/articles/PMC6968572/pdf/ZEPT_11_1708618.pdf

Haczkewicz, K. M., Shahid, S., Finnegan, H. A., Moninn, C., Cameron, C. D., & Gallant, N. L. (2024). Adverse childhood experiences (ACEs), resilience, and outcomes in older adulthood: A scoping review. Child Abuse & Neglect, 106864.

Hanlon, P., McCallum, M., Jani, B. D., McQueenie, R., Lee, D., & Mair, F. S. (2020). Association between childhood maltreatment and the prevalence and complexity of multimorbidity: A cross-sectional analysis of 157,357 UK Biobank participants. Journal of comorbidity, 10, 2235042X10944344. Retrieved from https://pmc.ncbi.nlm.nih.gov/articles/PMC7416137/pdf/10.1177_2235042X10944344.pdf

Hardcastle, K., & Bellis, M. (2018). Routine enquiry for history of adverse childhood experiences (ACEs) in the adult patient population in a general practice setting: A pathfinder study. Public Health Wales NHS Trust.

Hepsomali, P., & Groeger, J. A. (2021). Diet and general cognitive ability in the UK Biobank dataset. Scientific reports, 11(1), 11786. Retrieved from https://pmc.ncbi.nlm.nih.gov/articles/PMC8175590/pdf/41598_2021_Article_91259.pdf

Ho, F. K., Celis-Morales, C., Gray, S. R., Petermann-Rocha, F., Lyall, D., Mackay, D., . . . Pell, J. P. (2020). Child maltreatment and cardiovascular disease: quantifying mediation pathways using UK Biobank. BMC medicine, 18(1), 143. Retrieved from https://bmcmedicine.biomedcentral.com/counter/pdf/10.1186/s12916-020-01603-z.pdf

Holmberg, M. J., & Andersen, L. W. (2022). Collider bias. Jama, 327(13), 1282–1283. Retrieved from https://watermark.silverchair.com/jama_holmberg_2022_gm_220003_1648650493.36368.pdf?token=AQECAHi208BE49Ooan9kkhW_Ercy7Dm3ZL_9Cf3qfKAc485ysgAAA0AwggM8BgkqhkiG9w0BBwagggMtMIIDKQIBADCCAyIGCSqGSIb3DQEHATAeBglghkgBZQMEAS4wEQQM8tCz-G4IkSviu5BjAgEQgIIC8yYpg_o0aXTkjbilBcD3WQZ-9g7PccC2XYzICV62l-oMxauVWthkwekE4o5CzLXD80ab2vwXhlfg2tS9CQkaOvRkmL9NtwhqIw_rIENR_4kJSKtSTot5eslN5zoToFMvg2BN8Et_0sIustRl6eZOgKuYhvRvuuIRejWZApyvi4yLTN6t4gGK2foYlVuhegka5hciME1ES69koOW2gAOa3b7BANg6tA9jBvUgBODTALbvkG92dAvxwH9l8l32NNFr1ysiQdJ9OiUo5f4o8QmzdO6PFfNCaGdC1p1OsD9GDUQvEozKmwk7EOvul-ttQ2RQ0Ly-yk_UoHxozaN8-z01JvFKe-D61GPUssIRJEwxIBNS0F7t_GnR9Up_9ICb8I3inUwW_-nTg1Jwd8ipofauedVEm0Y6CBC2oW-asIbKNtZfEA8oh7xrB5-i2yvQQrktGlSt_mO1wJC6cGkHfWvcM1kJ9eH_dU-pIn2L_BZ8H_LtSDDTJ3HgFj_sxbKYg4YuKg-IJWDtq5DHYGkIhmczToudo9f38WunVwAVGx0Q7NKeV36IBbYrjYx2D1MtCnB8VjgOS1ab29cgFNPLtktwdq6SxIXI3SkHdemzpHOb05fT0QvneY6DSKQwBJ1WuaWpQ7dp_9Y1Alfja8dw9l7S3HtTFbebnd0bJRY_8nHjKYX63iUfkuYR4lS9RRBHZIh26fYyDTfNUJqaPo0k2INhvN9I3-oZCyaepRi0B-IkZKBAiCn50fZ6NXhJ6uCSB5CqOZ85EQWmmGIZlEpjQYF9HYZG1wKpMc9WaUtCMHXRbvxF1DuEwRRlkUDjtYw5-2NHmRujl6a_MXbiMrHIMbhZTBxU6KSAEMr6ilDgUYgvOPQN4DcQr7RLbvr7GtI3XI5QYdTdDFPVjeTkg9D8MCLedYm_ibSCUMWlPF6eh1rrufdAWh4skd9cOfXS9p83c2N8g08hyV4JxgofBj9FIphglZr0BS2nkdHJ4zyeQoEq-FN4moUo

Horwitz, A. V., Widom, C. S., McLaughlin, J., & White, H. R. (2001). The impact of childhood abuse and neglect on adult mental health: A prospective study. Journal of health and social behavior, 184–201.

Hughes, K., Bellis, M. A., Hardcastle, K. A., Sethi, D., Butchart, A., Mikton, C., . . . Dunne, M. P. (2017). The effect of multiple adverse childhood experiences on health: a systematic review and meta-analysis. The Lancet public health, 2(8), e356–e366. Retrieved from https://www.thelancet.com/pdfs/journals/lanpub/PIIS2468-2667(17)30118-4.pdf

Hughes, K., Ford, K., & Bellis, M. (2020). Adverse Childhood Experiences (ACEs) and Diabetes| A Brief Review.

Jackson, S. E., Kirschbaum, C., & Steptoe, A. (2017). Hair cortisol and adiposity in a population-based sample of 2,527 men and women aged 54 to 87 years. Obesity, 25(3), 539–544. Retrieved from https://pmc.ncbi.nlm.nih.gov/articles/PMC5324577/pdf/OBY-25-539.pdf

Jones, C. M., Merrick, M. T., & Houry, D. E. (2020). Identifying and preventing adverse childhood experiences: implications for clinical practice. Jama, 323(1).

Kalmakis, K. A., Meyer, J. S., Chiodo, L., & Leung, K. (2015). Adverse childhood experiences and chronic hypothalamic–pituitary–adrenal activity. Stress, 18(4), 446–450.

Katzourou, I. K., Barroso, I., Benger, L., Ingason, A., Stow, D., Tsang, R., . . . Owen, M. J. (2025). Contributions of common and rare genetic variation to different measures of mood and anxiety disorder in the UK Biobank. BJPsych Open, 11(3), e97. Retrieved from https://www.cambridge.org/core/services/aop-cambridge-core/content/view/71FF0C5A535ED2BD9063DC7D745ED538/S2056472425000432a.pdf/div-class-title-contributions-of-common-and-rare-genetic-variation-to-different-measures-of-mood-and-anxiety-disorder-in-the-uk-biobank-div.pdf

Katzourou, I. K., consortium, L., Barroso, I., Clayton, J., Khandaker, G., Stow, D., . . . Bree, M. B. M. v. d. (*in prep*). Neurodevelopmental copy number variants increase risk of internalising and cardiometabolic multimorbidity: findings from UK Biobank.

Lin, L., Wang, H. H., Lu, C., Chen, W., & Guo, V. Y. (2021). Adverse childhood experiences and subsequent chronic diseases among middle-aged or older adults in China and associations with demographic and socioeconomic characteristics. JAMA network open, 4(10), e2130143–e2130143. Retrieved from https://watermark.silverchair.com/lin_2021_oi_210873_1654876934.99549.pdf?token=AQECAHi208BE49Ooan9kkhW_Ercy7Dm3ZL_9Cf3qfKAc485ysgAAAy8wggMrBgkqhkiG9w0BBwagggMcMIIDGAIBADCCAxEGCSqGSIb3DQEHATAeBglghkgBZQMEAS4wEQQMjdA2jnIJK3v_Tv2IAgEQgIIC4nR3Kk1audivqW9odekOP5gw-KIm4WohDCRZphKkYGqrYRM6MKrj8RYL5t7WqpS6rdrr9b7yWUQDd9FzODYuS9k4BeVuSSzdLWpDk70lqy1Ox-zAf3UeelfotnrMaS5CS87ayqbOk0DUlKal6FBpXsQShDeWMWXBFH91ghiSuk8OAfRoBSxjZiifmBWeMeU-RCXx0rjXWZ78919ejNoNU8Z_-QsyoURB00OtNy5fIk_-qg1UtayHAbQj8kQ6bYlfwhv5sOWwkXebaXk2JyCg1xYDeSYq8aEbHJf3_XnS3GmzJfMKfiJ0tmMnBfljPoZTdLKcDdv5BYBr8e8-xXFqOID_0dLhKv8rQiR0X8l5fd6PYxs9OE5aUGK6YMWI2E950_iApiW3K4x6j9-VftXqiu84nKMLM3acQCSFg_VrhatniSmgcdXNbUlwzAd3SQ1CBeY2Wogm8NEiOo2dih_2Bpo7atHKMR8RtHjlFMGh8yMLO2U8gq_EH8HbN1eYvb0OA-YpbLgiFHyCwu5AnoJoD56w6usYkSusfWvdVUkT-SHjkyaAeGid5apceb010kD0IBd8Tsk0Lhfook5ZUf0ay2E0-yfMs1joBzYpiS0VyRGcLlqtAhw5g4GlrKakaZRprSemarUCL1l4U0UY_sd8Js8aZauJSvh7HmX4H0oKY_ZQp8HdEV2TpOIcCRJHvf9EJ77RJLgGFtUkgOA6HrgTu3Q8i43bGAKVqEJPt2hEWKKdr6HUhOamNLbvmxLUO5ZOBmMiktPDeHHK-ZXum5hGILf8jnFZjyWgIJJVobdm1d27rFasEhYJH47NVfklLHOh-sX5VCvteDek1IZkKgp2EZhuFWqMOKpCVHD1XRk023wZMkVxZDo6pkFRcJmf0yr7ToICa1AOMbdsgooT7_0wip9kJrWrYGXB09fmtYkCKvDL21Il84zkvApQ76JSoH5xKTLR_2CKPnv4skdZ0c792VHKKw

Loveday, S., Hall, T., Constable, L., Paton, K., Sanci, L., Goldfeld, S., & Hiscock, H. (2022). Screening for adverse childhood experiences in children: a systematic review. Pediatrics, 149(2), e2021051884. Retrieved from https://watermark.silverchair.com/peds_2021051884.pdf?token=AQECAHi208BE49Ooan9kkhW_Ercy7Dm3ZL_9Cf3qfKAc485ysgAAA1EwggNNBgkqhkiG9w0BBwagggM-MIIDOgIBADCCAzMGCSqGSIb3DQEHATAeBglghkgBZQMEAS4wEQQMQCDirA1Nr0Q0q2OOAgEQgIIDBCsdvPni6PJepxz4fUzXbcjQZzdlrAjJLvci3LT1XZmBSPymY2lVDClj34r7pSdG1g2aYE2RMwlf6WMz7ypFRW9E6Dnv1T08C9KKpVbkTYqxLhftE5pEzyCM9FXXL_aOW74M5G_3MrKkiRTpKEES6hMlRH6vx7T85CT0SneZvJkOFNDztiO-TXiHBOK-qacKJiXKgPV2zLAC2_YKv36ZiXe6ypK_gABrZO3nUBjyoXlWoD0UCxC-Wrdd_aCkTtFTYfQldPciXEReXgW5wRGly8YkretPk2EeBO5ukdCQkyw6aGEDDP7bpvJtxExKn6ba_27YdbSGwXuoBX8LK_W14kqc27zUttY9W-FEVVxJ2AFG1wDacrUrbdKKlIzfqbZ6XMVsvqhy5DNoKdPT7OK-iIkYByMO-7ay8pfNqdYoP6jlu-kY7I_-TWvAbCcggMKDLwjT7fAYDKpKvzaI0x0FtA0yh6Qznqyt3JDd_z4s9WeZ255wiH7GEfx6gEmIezeRp0d2qXujLwBU5P5xxvMu246AOnDco0Nkwa62GEM5gGbs7Jnny6tpING5Wlwn26FGPAndYR2s7CJA3OfgSzaL5AJocrDH6clGDywuVYRhZm06GxeOIMSEnjW4mRopTsrw-CpJHghZfjZqSSYW6KFLvJurMXCs0tVsWO4LmBM7HDjmOmFmN-2qeKdZdoQH8xqxPUyCvj35QDVkKV_Tsdx25ezw0SqWYG8fSnoR-LATxSX9lyKgzvNxvv6dOEbJzZoLfLH5nAJkIf9vveXSeJKurqaHztN4HUdnp48W_9QIh2NoKt0D2BKiroyL1MvXl6Qhv2WZwNRjH4tBvj5KOILVkRQpsihnWyFzrQWxry4kMhVsJSm87nQQx7_b140zxetfcsB_aP0n3M3C_vyGkhO5frqQISdN3Ove4kCOEZVI1uajDFDyUyU3onUxyNk_oSTF0cx3DH1DJoY0Wn7tqofJuQVg2aF3J8HABg5qLtMOxPJ8GDagmihI2pQ0DErkPsKZYmGMyQs

Madden, J. M., Lakoma, M. D., Rusinak, D., Lu, C. Y., & Soumerai, S. B. (2016). Missing clinical and behavioral health data in a large electronic health record (EHR) system. Journal of the American Medical Informatics Association, 23(6), 1143–1149. Retrieved from https://watermark.silverchair.com/ocw021.pdf?token=AQECAHi208BE49Ooan9kkhW_Ercy7Dm3ZL_9Cf3qfKAc485ysgAAA1wwggNYBgkqhkiG9w0BBwagggNJMIIDRQIBADCCAz4GCSqGSIb3DQEHATAeBglghkgBZQMEAS4wEQQMWgreeV05uzYcrM7vAgEQgIIDD6LZLxvrtubQ0MOwScykdNqSRof7BTFR7eZaPqxvbIz9ZGcilB-pHzZEPFAZrhbOZ3d0l5Ldf-6buIeWZQf2Q_3EAdoAeeJCFhPHzgN1KhWKuMTC8Y-oZvqwWP-1CaqtNZQRHvUz62aTCz2a5KdXgWwoWcMX2z7zlShspNl5r74LRMwrrZm6cm2TuPUu1tFQu2uY7Ck5OaPHEgA_UhbQRIy3r33fwq6xjL8ZuJI3yeESs4vMNfbXHFSYyyT-rQf6z5fXL8_dWgJHYKW4pM0l9fQFv0H-FWCFGSTg6hopyzGbF1KHplgUytB7e6oqVgugNydK04STWThz1pZUQ7q81qCo5OGs4IwSrRZq-EEpfbFW4awYL2S9c9Klqo2ZxMysjOneB9F3rGripK49RzSHuNi9BENVrLS20P25imvhaZbNOqxj97IuFnXKyhxfSf02ehu3aBjc5IZZMxYiJH1umBLIA5YTF1V2OPNUx70ttlzddvztTx7NdQ9s0--F1qopxklePbTnzXeZD09sLtQEUp47G8saHXVcemaFGfQagSFySFq_frmQy8AqZ7FcKnXEVsyoLTB4LzMhsl6a_PAIG5Z13l0v9qns6SStuyVQ4TaG9bQ8hm7SvSoDRHPQOJ4EiJD_L0E_AgJv0Pu2d0IZvyYu8wT7Ckm_p96PgkCEwe1UKBYDWQcA2vF1ndzUgXmGDgq4lldqIEHS1l4Eu-5SzK6_HwpeSS3VoSqtTxAtsDGWXddg3KsxZfTltrhaAzLBDvM2-ivbKaadIee4xsc2FhLw-gdG34g5-QajoUco_yJ_WX6Qr5864to4n-BWE-QP9T73_AOsnjazyY4Rt8y3bWePc12y4TEtQ_sAKY7UDv4QPqfYRp_Gpj8JvNE5VhypgfsdEgy1jS8s7jD79TtSOApio67h5NDjBvHvFlP-SVM-5TXbdc2qRt1dJrqZAGQ4yw3Y477bmVwWboCddic6dtitcg6IsZ28QgCkP6aj5GXfbKgNIMGRu5GoIN6EvZnElaEmGWEnF9fzjB_bDXTUg

Masood, B., & Moorthy, M. (2023). Causes of obesity: a review. Clinical Medicine, 23(4), 284–291. Retrieved from https://pmc.ncbi.nlm.nih.gov/articles/PMC10541056/

McGill Jr, H. C., McMahan, C. A., Zieske, A. W., Tracy, R. E., Malcom, G. T., Herderick, E. E., & Strong, J. P. (2000). Association of coronary heart disease risk factors with microscopic qualities of coronary atherosclerosis in youth. Circulation, 102(4), 374–379.

McLean, C. P., Asnaani, A., Litz, B. T., & Hofmann, S. G. (2011). Gender differences in anxiety disorders: prevalence, course of illness, comorbidity and burden of illness. Journal of psychiatric research, 45(8), 1027–1035.

McLennan, J. D., Gonzalez, A., MacMillan, H. L., & Afifi, T. O. (2024). Routine screening for adverse childhood experiences (ACEs) still doesn’t make sense. Child Abuse & Neglect, 106708.

Meloni, A., Cadeddu, C., Cugusi, L., Donataccio, M. P., Deidda, M., Sciomer, S., . . . Mercuro, G. (2023). Gender differences and cardiometabolic risk: the importance of the risk factors. International journal of molecular sciences, 24(2), 1588. Retrieved from https://pmc.ncbi.nlm.nih.gov/articles/PMC9864423/pdf/ijms-24-01588.pdf

Mercer, S., Furler, J., Moffat, K., Fischbacher-Smith, D., & Sanci, L. (2016). Multimorbidity: technical series on safer primary care: World Health Organization.

Mercer, S. W., Gunn, J., Bower, P., Wyke, S., & Guthrie, B. (2012). Managing patients with mental and physical multimorbidity. In (Vol. 345): British Medical Journal Publishing Group.

Merrick, M. T. (2019). Vital signs: estimated proportion of adult health problems attributable to adverse childhood experiences and implications for prevention—25 states, 2015–2017. MMWR. Morbidity and mortality weekly report, *68*.

Merrick, M. T., Ports, K. A., Ford, D. C., Afifi, T. O., Gershoff, E. T., & Grogan-Kaylor, A. (2017). Unpacking the impact of adverse childhood experiences on adult mental health. Child Abuse & Neglect, 69, 10–19.

Mersky, J. P., Choi, C., Lee, C. P., & Janczewski, C. E. (2021). Disparities in adverse childhood experiences by race/ethnicity, gender, and economic status: Intersectional analysis of a nationally representative sample. Child Abuse & Neglect, 117, 105066.

Metzler, M., Merrick, M. T., Klevens, J., Ports, K. A., & Ford, D. C. (2017). Adverse childhood experiences and life opportunities: Shifting the narrative. Children and youth services review, 72, 141–149. Retrieved from https://pmc.ncbi.nlm.nih.gov/articles/PMC10642285/pdf/nihms-1931034.pdf

Moore, S. A., & Zoellner, L. A. (2007). Overgeneral autobiographical memory and traumatic events: an evaluative review. Psychological bulletin, 133(3), 419. Retrieved from https://pmc.ncbi.nlm.nih.gov/articles/PMC2665927/

Obi, I. E., McPherson, K. C., & Pollock, J. S. (2019). Childhood adversity and mechanistic links to hypertension risk in adulthood. British journal of pharmacology, 176(12), 1932–1950. Retrieved from https://pmc.ncbi.nlm.nih.gov/articles/PMC6534788/pdf/BPH-176-1932.pdf

Office for National Statistics. (2020a). Child abuse in England and Wales: March 2020. . Retrieved from https://www.ons.gov.uk/peoplepopulationandcommunity/crimeandjustice/bulletins/childabuseinenglandandwales/march2020

Office for National Statistics. (2020b). Child physical abuse in England and Wales: year ending March 2019. Retrieved from https://www.ons.gov.uk/peoplepopulationandcommunity/crimeandjustice/bulletins/childabuseinenglandandwales/march2020

Piccinelli, M., & Wilkinson, G. (2000). Gender differences in depression: Critical review. The British Journal of Psychiatry, 177(6), 486–492.

Shi, W., Huang, Y., & Jin, C. (2022). Association of adverse childhood experiences with subsequent kidney disease among middle-aged and older adults in China: A national analysis. medRxiv, 2022.2006. 2008.22276145.

Soley-Bori, M., Ashworth, M., Bisquera, A., Dodhia, H., Lynch, R., Wang, Y., & Fox-Rushby, J. (2021). Impact of multimorbidity on healthcare costs and utilisation: a systematic review of the UK literature. British Journal of General Practice, 71(702), e39–e46. Retrieved from https://pmc.ncbi.nlm.nih.gov/articles/PMC7716874/pdf/bjgpjan-2021-71-702-e39-oa.pdf

Stephens, A., Allardyce, J., Weavers, B., Lennon, J., Jones, R. B., Powell, V., . . . Osborn, D. (2023). Developing and validating a prediction model of adolescent major depressive disorder in the offspring of depressed parents. Journal of child psychology and psychiatry, 64(3), 367–375.

Steptoe, A., & Frank, P. (2023). Obesity and psychological distress. Philosophical Transactions of the Royal Society B, 378(1888), 20220225. Retrieved from https://pmc.ncbi.nlm.nih.gov/articles/PMC10475872/

Stokes, J., Guthrie, B., Mercer, S. W., Rice, N., & Sutton, M. (2021). Multimorbidity combinations, costs of hospital care and potentially preventable emergency admissions in England: A cohort study. PLoS medicine, 18(1), e1003514. Retrieved from https://pmc.ncbi.nlm.nih.gov/articles/PMC7815339/pdf/pmed.1003514.pdf

Suglia, S. F., Koenen, K. C., Boynton-Jarrett, R., Chan, P. S., Clark, C. J., Danese, A., . . . Isasi, C. R. (2018). Childhood and adolescent adversity and cardiometabolic outcomes: a scientific statement from the American Heart Association. Circulation, 137(5), e15–e28. Retrieved from https://pmc.ncbi.nlm.nih.gov/articles/PMC7792566/pdf/nihms-1587741.pdf

Sulieman, L., Cronin, R. M., Carroll, R. J., Natarajan, K., Marginean, K., Mapes, B., . . . Ramirez, A. (2022). Comparing medical history data derived from electronic health records and survey answers in the All of Us Research Program. Journal of the American Medical Informatics Association, 29(7), 1131–1141. Retrieved from https://watermark.silverchair.com/ocac046.pdf?token=AQECAHi208BE49Ooan9kkhW_Ercy7Dm3ZL_9Cf3qfKAc485ysgAAA14wggNaBgkqhkiG9w0BBwagggNLMIIDRwIBADCCA0AGCSqGSIb3DQEHATAeBglghkgBZQMEAS4wEQQMqifKs_D1_jANOnifAgEQgIIDEVUTLKz75OpS4BRf72I5kHA1QivQi6wj3QL4mB_grjda43GNqBXumc2rrCgJgANWLX6GaKhyjU9-mqGiyQSNAjBEpxDeXuBWWoVYuvfqkBWgT5ISDT3XwepptklqO5UBKR-sJYb9popsFAjVjeVxkwELbQkB5kEiZEm-X1E8IjTe_y4pS_BujpztdI5lH_Nf0fv-uDfX4ZHOGS_woWNjlEw5jYK7D5A25O6xG5P6PwQXveztN2TQdFQndm5k9f4HdbmizEYA5oQB-yqqtJqjqhdSOYgEBn9oAbs_vvYbny8na-G-bMtG_fR_lvjt7u4dg1XirQt3lfMYARxMI-5udleIXuVBQDGYfCZ3bfCsUNinS5G8VQKWR1Pzb898ROOZggWH9Kz-iMxcvCqSr6dzRgZqSQ_2ppUieKNq_Bob2nZo4qejQnKdTobvrpvqqKg7c4MvvuN0IEYERGZgGxfiE7266PDpEVP8NxKCfWyS4djjVYN3Jj422LLiAx2XiynD-wiG6xkiAAVg-spf-58aQ0nGzuwWKpwVb_xGsdU5VznMnEzfnXvVYzIeQwzd-Hyg36qtI5dFK1GuK_Td5yiJWgbBpON1IJXpooPKQSSHeguLCcNLr5sHsjk6wKUnEHpxass2sP3foBvQRFf-dz8Firi-0oUpHRPzKm-D8X4dmXvKq8tX-D0SLOPnjfT62xgFsN47uEKPdHofd9UJ7vCybXNcanYDsvIwBwF002BKrhD9m84nFmfrW0BaM-fIT5v-CJ2ApaShoHRPA_bgCDrP8VyzjujP3-dUXZOO97O2fFHPeI5ctC1sG6mCMGXxGYJPFrkQOyGMRnSrm0DY5RR8MtqxeRZ8sxwwvi6og1rIrYj5Bc2I4GOF4j2zM_HmzZI1nL0e0OXdeR7LJLh3krHZLvHiYhD7oxMYIfcjJGb12iyKy7BbvGKU_GhzySRM_2yYA3XusyH1nGcbwK78laNdekVyYVb6wGcurlG9Tcg7LQeBg9R-Fh_3syYYDTNBlxmlPVjxvsntl0yw-duX8a1OmtkG

Taylor, K., & Demakakos, P. (2024). Adverse childhood experiences and trajectories of multimorbidity in individuals aged over 50: evidence from the English Longitudinal Study of Ageing. Child Abuse & Neglect, 149, 106653.

Testa, A., Zhang, L., Jackson, D. B., Ganson, K. T., Raney, J. H., & Nagata, J. M. (2024). Adverse childhood experiences and unhealthy dietary behaviours in adulthood. Public health nutrition, 27(1), e40. Retrieved from https://pmc.ncbi.nlm.nih.gov/articles/PMC10882537/pdf/S1368980024000144a.pdf

The World Health Organisation. (2023a). Anxiety disorders. Retrieved from https://www.who.int/news-room/fact-sheets/detail/anxiety-disorders

The World Health Organisation. (2023b). Depressive disorder (depression). Retrieved from https://www.who.int/news-room/fact-sheets/detail/depression

Torres, S. J., & Nowson, C. A. (2007). Relationship between stress, eating behavior, and obesity. Nutrition, 23(11-12), 887–894.

UK Biobank. (2024). Health-related outcomes data. Retrieved from https://www.ukbiobank.ac.uk/enable-your-research/about-our-data/health-related-outcomes-data

Walsh, D., McCartney, G., Smith, M., & Armour, G. (2019). Relationship between childhood socioeconomic position and adverse childhood experiences (ACEs): a systematic review. J Epidemiol Community Health, 73(12), 1087–1093. Retrieved from https://pmc.ncbi.nlm.nih.gov/articles/PMC6872440/pdf/jech-2019-212738.pdf

Wang, M., Zhou, T., Song, Q., Ma, H., Hu, Y., Heianza, Y., & Qi, L. (2022). Ambient air pollution, healthy diet and vegetable intakes, and mortality: a prospective UK Biobank study. International journal of epidemiology, 51(4), 1243–1253. Retrieved from https://pmc.ncbi.nlm.nih.gov/articles/PMC9365625/pdf/dyac022.pdf

Zhen-Duan, J., Colombo, D., Cruz-Gonzalez, M. A., Hoyos, M., & Alvarez, K. (2023). Adverse childhood experiences and alcohol use and misuse: Testing the impact of traditional and expanded adverse childhood experiences among racially/ethnically diverse youth transitioning into adulthood. Psychological trauma: theory, research, practice, and policy, 15(S1), S55.

Zulman, D. M., Haverfield, M. C., Shaw, J. G., Brown-Johnson, C. G., Schwartz, R., Tierney, A. A., . . . Israni, S. T. (2020). Practices to foster physician presence and connection with patients in the clinical encounter. Jama, 323(1), 70–81. Retrieved from https://watermark.silverchair.com/jama_zulman_2020_sc_190007.pdf?token=AQECAHi208BE49Ooan9kkhW_Ercy7Dm3ZL_9Cf3qfKAc485ysgAAAy0wggMpBgkqhkiG9w0BBwagggMaMIIDFgIBADCCAw8GCSqGSIb3DQEHATAeBglghkgBZQMEAS4wEQQMdMN2IbIPijxjEC0SAgEQgIIC4JuWXQS0QRnZ9jJ7XyJyD52QoGnZj2vaayIL90RwyvSFEkp1VjaC4BDh6XBLQgs16laoFCZ7ctmRCYyYGEazICdwrOySsRQYrdyQVqJkJ77bxTwgxZN4RilPECdnPrpQh_oXfGfWscrNsVWYoa08WV0kzsoWywn1mdjO1iBaqVqfcyQNy8oBw6zTIjQK1KDcvcLxa1uQV66r2YzNt1gU-mU0NTpxzteZT3jr8G_q5KOE89OrV2IEUmdnDo9YCcZ15w6I_jluXyScg6djjJhiWL0m-WMP1ksFVLQyNKHMmB5yIWlw3LDAW5gZA283HftTJoEUmCscK3uV5VCIRivOPnwXhCfMYPGqBf0DfrIcaLbrKqC6NGuyCfPVt3Y0pe-dHwf0UW_fvNvVretmy6kcBieMc-pmuCySovduVxQ2wxzX5f3xu4KnVA_Jz90VoqQxYFXRFBJvDCp5518wemljYwYl4_3oHylDPibTgfNSIFyLddTRmXZtF4xhWuQgCaYxg-1ra2bA8_pkJGI7qGb1CWkv_0eKf-8Gk0kblgd29qFaEpSv_LHpGWrJ3gQM_qz8GD-PIvJ00sopKuRhjOM9o0g1b7DmBZj9Hiq_EfjO3CFqzBhtyIfP3lOl4jb29Q5H5LccmEZw79E_iQlHYTzebanNz8_n5wtEPJXqgQybA5hVTThWOSDetvC2FVb_j6pnsPRLsXshyRwRy_ie0iJjrhPgAc0zMauJHYrijEn99AyK4GrJDTyXYlpvdr-NTmClyDGuENamKtKolT59d788Aqbyl5stc0I_OTWhd1Rr8-8_QhVA3d4XOvtlLFefbACmtAjCz3lQFMb2FjToztIDtWKDNMCXcbt5mcdfQvW6W1wJs13NJ5TalSs138Cmz5p57MF8SS_weuFRkzrmiE4cvAexh28T0ctRYP-C5kZGx039pN6CeqtLIubyJPXY4-d9Bwj4vAhjT--gyP5Wabuy30w

