## Supplementary material for "Adverse childhood experiences and multimorbidity of internalising and cardiometabolic conditions in mid to older age"

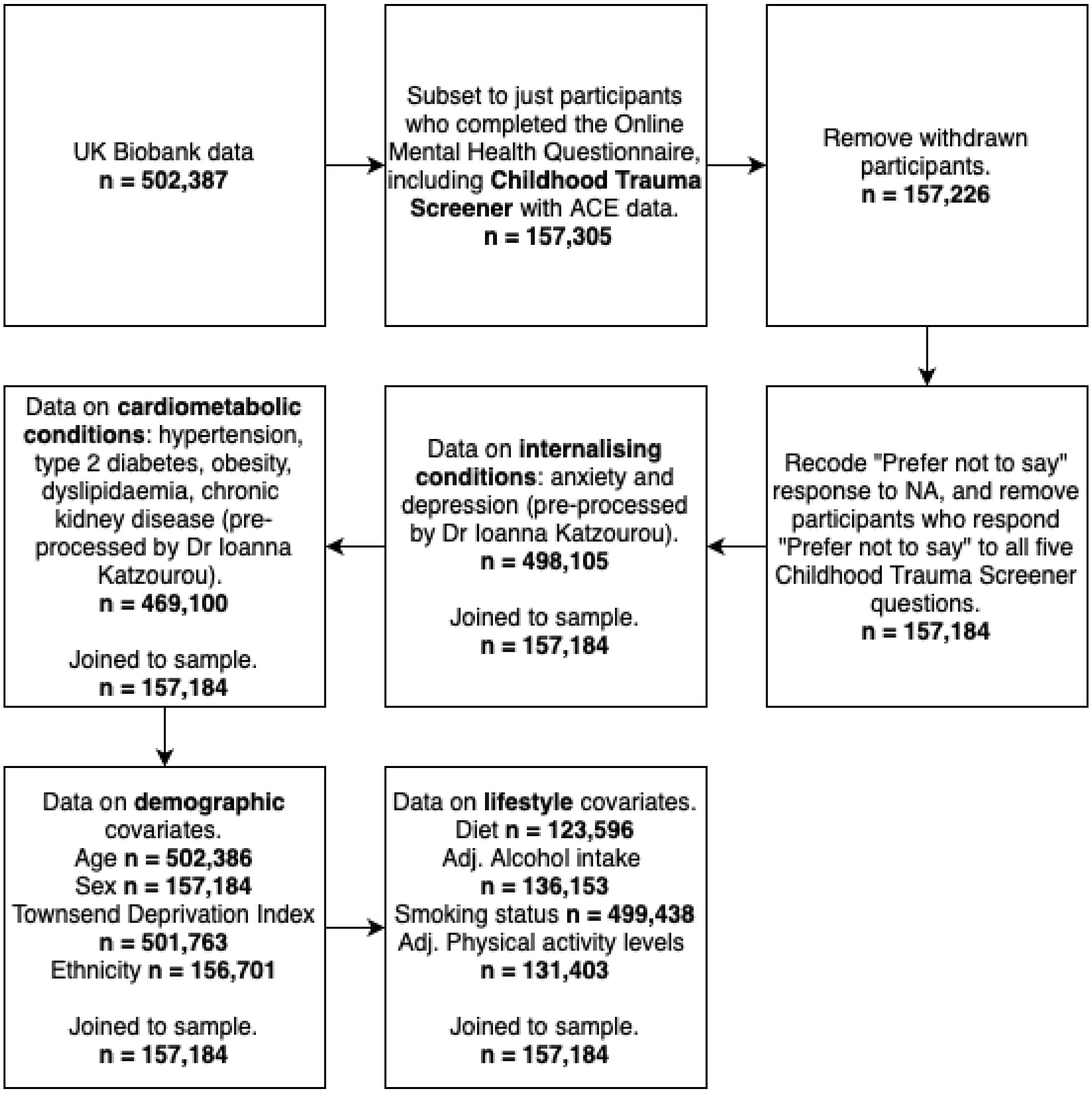


*Supplementary Figure 1*. Flow chart of data cleaning in R, including the exclusions to base sample and corresponding sample sizes based on availability of adverse childhood experiences data. For sample sizes of individual covariates after joining to base sample, see *Table 1.*

| **Adverse childhood experience** | **Childhood Trauma Screener question** | **Threshold for dichotomisation (as in Ho et al., 2020)** |
|---|---|---|
| Emotional neglect | When I was growing up, I felt loved | **Never true, Rarely true, Sometimes true = 1**; Often, Very often true = 0 (reverse coded) |
| Physical abuse | When I was growing up, people in my family hit me so hard that it left me with bruises or marks | Never true, Rarely true = 0; **Sometimes true, Often, Very often true = 1** |
| Emotional abuse | When I was growing up, I felt that someone in my family hated me | Never true, Rarely true = 0; **Sometimes true, Often, Very often true = 1** |
| Sexual abuse | When I was growing up, someone molested me (sexually) | Never true = 0; **Rarely true, Sometimes true, Often, Very often true = 1** |
| Physical neglect | When I was growing up, there was someone to take me to the doctor if I needed it | **Never true, Rarely true, Sometimes true = 1**; Often, Very often true = 0 (reverse coded) |
| Possible responses to the 5 questions were "Very often true", "Often", "Sometimes true", "Rarely true", "Never true" and "Prefer not to say".<br>Responses were recoded numerically from 1-5 with 5 denoting more severe trauma. The two neglect variables were reverse coded for consistency with the abuse variables (e.g. a response of "Never true" to the physical neglect question was coded 5, versus a response of "Very often true" to the physical abuse question was coded 5). | | |

*Supplementary Table 1.* Details of adverse childhood experience type, questions as in Childhood Trauma Screener and the threshold for dichotomisation where '0' is absent and '1' is present. Bold text highlights the question responses indicating the presence of each ACE.

| Derived variable | Coding | |
|---|---|---|
| | Present ‘1’ | Absent ‘0’ |
| **Any IC** | Diagnosis of either depression and/or anxiety. | Neither depression nor anxiety. |
| **Any CMC** | Diagnosis of either hypertension, T2D, obesity, CKD and/or dyslipidaemia. | No diagnosis of hypertension, T2D, obesity, CKD or dyslipidaemia. |
| **Any IC plus hypertension** | Diagnosis of either depression and/or anxiety and hypertension. | Neither any IC nor hypertension. |
| **Any IC plus T2D** | Diagnosis of either depression and/or anxiety and type 2 diabetes. | Neither any IC nor type 2 diabetes. |
| **Any IC plus obesity** | Diagnosis of either depression and/or anxiety and obesity. | Neither any IC nor obesity. |
| **Any IC plus CKD** | Diagnosis of either depression and/or anxiety and chronic kidney disease. | Neither any IC nor chronic kidney disease. |
| **Any IC plus dyslipidaemia** | Diagnosis of either depression and/or anxiety and dyslipidaemia. | Neither any IC nor dyslipidaemia. |
| **Any ICM-MM** | Diagnosis of any IC and any CMC (as above). | Neither any IC plus any CMC, nor any IC or any CMC alone. |

*Supplementary Table 2.* Details of coding of individual and multimorbid health presentations, where ‘1’ is present and ‘0’ is absent.

### Further information on demographic variables

All demographic data was taken at recruitment. We used the UK Biobank variable “Sex” but referred to it as “gender” throughout the work because the data are not indicative of biological sex inferred by genetic data. The reference category for gender was female ‘1’. SES was measured using the Townsend Deprivation Index (TDI), comprising four variables: overcrowding, unemployment, non-home ownership and non-car ownership. A score of zero indicates the average material value of an area, positive values indicate high material deprivation and negative values indicate relative affluence. Ethnicity was categorised as white and non-white.

**Further information on lifestyle variables**

All lifestyle variables were taken from the touchscreen questionnaire completed at recruitment. Participants stated their "Alcohol drinker status" as "current", "previously" or "never". Those answering "current" or "previous" reported their "Alcohol intake frequency". Those reporting drinking one to three times a month or less estimated the number of glasses of alcohol consumed in an average month. Those reporting more frequent alcohol consumption estimated the number of glasses of alcohol consumed in an average week. Glasses per month were converted to glasses per week. Based on the UK Biobank defined units (Daviet et al. 2022), units of alcohol consumed per week were calculated for all participants. Units per week of over 100 were considered to be high pathological levels indicating individuals outwith the sample of interest and were coded as missing.

A healthy diet score (ranging from '0' least healthy to '7' most healthy) was derived using data collected on consumption of 7 food groups - fruits, vegetables, fish, unprocessed meat, processed meat, wholegrains and refined grains, as is common in previous literature (Hepsomali and Groeger 2021; Wang et al. 2022). Participants stated the frequency of consumption of each food group. Cut off scores indicating "healthy" and "unhealthy" serving sizes of each food group were validated in (Liu et al. 2023), e.g. four or more servings of fruit (dried and fresh) a day was considered a healthy intake goal. Participants meeting this cut-off would receive a score of '1' for this food group. The healthy diet score was calculated by combining scores for all 7 food groups.

The Metabolic Equivalent of Task (MET) is a measurement of how much energy is used during activities. One MET is the amount of energy used when a person sits quietly and acts as a point of reference for other physical activities (Byrne et al. 2005). Total MET in minutes per week for all types of activity (walking, moderate, vigorous) was converted into hours per week. Scores of over 168 hours per week were deemed implausible and coded as missing.

| | Combined | Females | Males | p-value[2] |
|---|---|---|---|---|
| Overall sample (%) | 157184 (100) | 88998 (56.62) | 68186 (43.38) | |
| **No ACEs (%)** | 104304 (66.36) | 57742 (64.88) | 46562 (68.29) | |
| **Any ACE[1] (%)** | 52880 (33.64) | 31256 (35.12) | 21624 (31.71) | **$< 2.2 \times 10^{-16}$** |
| **1 ACE (%)** | 32151 (20.45) | 17925 (20.14) | 14226 (20.86) | |
| **2 ACEs (%)** | 12233 (7.78) | 7506 (8.43) | 4727 (6.93) | |
| **≥3 ACEs (%)** | 8496 (5.41) | 5825 (6.55) | 2671 (3.92) | |
| **Emotional Neglect (%)** | 35247(22.51) | 20530 (23.15) | 14717 (21.67) | **$< 2.2 \times 10^{-16}$** |
| *NA (%)* | *585 (0.37)* | *320 (0.36)* | *265 (0.39)* | |
| **Physical Neglect (%)** | 8991 (5.76) | 5429 (6.15) | 3562 (5.25) | **$< 2.2 \times 10^{-16}$** |
| *NA (%)* | *1030 (0.66)* | *655 (0.74)* | *375 (0.55)* | |
| **Emotional Abuse (%)** | 14978 (9.56) | 10257 (11.56) | 4721 (6.94) | **$< 2.2 \times 10^{-16}$** |
| *NA (%)* | *410 (0.26)* | *275 (0.31)* | *135 (0.20)* | |
| **Physical Abuse (%)** | 12928 (8.24) | 7225 (8.14) | 5703 (8.38) | $8.52 \times 10^{-02}$ |
| *NA (%)* | *315 (0.20)* | *202 (0.23)* | *113 (0.17)* | |
| **Sexual Abuse (%)** | 13623 (8.77) | 9680 (11.05) | 3943 (5.82) | **$< 2.2 \times 10^{-16}$** |
| *NA (%)* | *1807 (1.15)* | *1404 (1.58)* | *403 (0.59)* | |

*Supplementary Table 3.* Count and prevalence (%) of each type of ACE in the full sample and split by gender. [1]Any ACE refers to reporting at least one of the following: emotional neglect, physical neglect, emotional abuse, physical abuse, sexual abuse. [2]P-values of binomial logistic regression between gender and types of ACE are reported. All p-values are corrected for multiple testing using the Bonferroni Correction, raw p-values are reported and those formatted in bold survived correction for multiple testing. *NA* indicates missing data.

| | Emotional Neglect | | | | | | | |
|---|---|---|---|---|---|---|---|---|
| | Odds ratio (95% CI) | p-value | Odds ratio (95% CI) | p-value | Odds ratio (95% CI) | p-value | Odds ratio (95% CI) | p-value |
| | Model 1 Unadjusted | | Model 2 Adjusted for demographic variables | | Model 3 Adjusted for lifestyle variables | | Model 4 Adjusted for demographic and lifestyle variables | |
| **Any IC[1]** | 1.90 (1.84-1.96) | **$< 2.2 \times 10^{-16}$** | 1.87 (1.81-1.93) | **$< 2.2 \times 10^{-16}$** | 1.85 (1.78-1.93) | **$< 2.2 \times 10^{-16}$** | 1.84 (1.76-1.92) | **$< 2.2 \times 10^{-16}$** |
| **Hypertension** | 1.08 (1.06-1.11) | **$1.15 \times 10^{-09}$** | 1.15 (1.12-1.18) | **$< 2.2 \times 10^{-16}$** | 1.08 (1.04-1.12) | **$3.95 \times 10^{-05}$** | 1.14 (1.10-1.18) | **$1.63 \times 10^{-11}$** |
| **T2D** | 1.25 (1.19-1.31) | **$< 2.2 \times 10^{-16}$** | 1.25 (1.19-1.32) | **$< 2.2 \times 10^{-16}$** | 1.21 (1.12-1.29) | **$1.31 \times 10^{-07}$** | 1.23 (1.15-1.32) | **$8.97 \times 10^{-09}$** |
| **Obesity** | 1.37 (1.31-1.42) | **$< 2.2 \times 10^{-16}$** | 1.34 (1.28-1.39) | **$< 2.2 \times 10^{-16}$** | 1.34 (1.26-1.42) | **$< 2.2 \times 10^{-16}$** | 1.33 (1.25-1.41) | **$< 2.2 \times 10^{-16}$** |
| **CKD** | 1.09 (1.03-1.15) | $4.21 \times 10^{-03}$ | 1.14 (1.08-1.21) | **$9.17 \times 10^{-06}$** | 1.04 (0.96-1.13) | 0.32 | 1.09 (1.00-1.18) | $3.81 \times 10^{-02}$ |
| **Dyslipidaemia** | 1.11 (1.07-1.14) | **$8.54 \times 10^{-11}$** | 1.15 (1.12-1.19) | **$< 2.2 \times 10^{-16}$** | 1.10 (1.05-1.14) | **$1.29 \times 10^{-05}$** | 1.14 (1.10-1.20) | **$1.05 \times 10^{-09}$** |
| **Any CMC[2]** | 1.13 (1.11-1.16) | **$< 2.2 \times 10^{-16}$** | 1.20 (1.17-1.23) | **$< 2.2 \times 10^{-16}$** | 1.13 (1.09-1.17) | **$< 2.2 \times 10^{-16}$** | 1.19 (1.15-1.24) | **$< 2.2 \times 10^{-16}$** |
| **Any IC plus hypertension** | 1.76 (1.67-1.84) | **$< 2.2 \times 10^{-16}$** | 1.77 (1.68-1.85) | **$< 2.2 \times 10^{-16}$** | 1.70 (1.59-1.81) | **$< 2.2 \times 10^{-16}$** | 1.72 (1.60-1.83) | **$< 2.2 \times 10^{-16}$** |
| **Any IC plus T2D** | 1.96 (1.79-2.15) | **$< 2.2 \times 10^{-16}$** | 1.89 (1.72-2.08) | **$< 2.2 \times 10^{-16}$** | 1.87 (1.63-2.14) | **$< 2.2 \times 10^{-16}$** | 1.86 (1.62-2.13) | **$< 2.2 \times 10^{-16}$** |
| **Any IC plus obesity** | 2.00 (1.86-2.14) | **$< 2.2 \times 10^{-16}$** | 1.89 (1.76-2.03) | **$< 2.2 \times 10^{-16}$** | 1.95 (1.75-2.16) | **$< 2.2 \times 10^{-16}$** | 1.88 (1.70-2.09) | **$< 2.2 \times 10^{-16}$** |
| **Any IC plus CKD** | 1.78 (1.60-1.97) | **$< 2.2 \times 10^{-16}$** | 1.81 (1.62-2.01) | **$< 2.2 \times 10^{-16}$** | 1.58 (1.36-1.85) | **$4.30 \times 10^{-09}$** | 1.61 (1.38-1.88) | **$1.66 \times 10^{-09}$** |
| **Any IC plus dyslipidaemia** | 1.75 (1.66-1.85) | **$< 2.2 \times 10^{-16}$** | 1.75 (1.66-1.86) | **$< 2.2 \times 10^{-16}$** | 1.78 (1.69-1.88) | **$5.93 \times 10^{-45}$** | 1.74 (1.61-1.88) | **$< 2.2 \times 10^{-16}$** |
| **Any ICM-MM[3]** | 1.83 (1.75-1.90) | **$< 2.2 \times 10^{-16}$** | 1.81 (1.74-1.89) | **$< 2.2 \times 10^{-16}$** | 1.78 (1.69-1.88) | **$8.19 \times 10^{-93}$** | 1.78 (1.69-1.89) | **$< 2.2 \times 10^{-16}$** |

*Supplementary Table 4.* Results of binary logistic regression models comparing the frequency of health outcomes in individuals with emotional neglect versus those without. [1]Any IC refers to a diagnosis of depression and/or anxiety from primary or secondary care records (*Supplementary Table 2*). [2]Any CMC refers to a diagnosis of hypertension, T2D, obesity, CKD and/or dyslipidaemia from primary or secondary care records (*Supplementary Table 2*). [3]ICM-MM refers to a diagnosis of any IC plus any CMC from primary or secondary care records. CKD refers to chronic kidney disease (*Supplementary Table 2*). Model 1 is not adjusted for any covariates. Model 2 is adjusted for demographic variables: age, gender, ethnicity and socioeconomic status. Model 3 is adjusted for lifestyle variables: diet, alcohol intake, smoking status and physical activity levels. Model 4 is adjusted for demographic and lifestyle variables: age, gender, ethnicity, socioeconomic status, diet, alcohol intake, smoking status and physical activity levels. All p-values are corrected for multiple testing using the Bonferroni Correction, raw p-values are reported and those formatted in bold survived correction for multiple testing.

| | Physical Neglect | | | | | | | |
|---|---|---|---|---|---|---|---|---|
| | **Odds ratio (95% CI)** | **p-value** | **Odds ratio (95% CI)** | **p-value** | **Odds ratio (95% CI)** | **p-value** | **Odds ratio (95% CI)** | **p-value** |
| | Model 1<br>Unadjusted | | Model 2<br>Adjusted for demographic variables | | Model 3<br>Adjusted for lifestyle variables | | Model 4<br>Adjusted for demographic and lifestyle variables | |
| **Any IC[1]** | 1.67 (1.59-1.76) | **$< 2.2 \times 10^{-16}$** | 1.67 (1.58-1.76) | **$< 2.2 \times 10^{-16}$** | 1.56 (1.45-1.69) | **$< 2.2 \times 10^{-16}$** | 1.58 (1.47-1.71) | **$< 2.2 \times 10^{-16}$** |
| **Hypertension** | 1.41 (1.35-1.48) | **$< 2.2 \times 10^{-16}$** | 1.34 (1.27-1.40) | **$< 2.2 \times 10^{-16}$** | 1.48 (1.39-1.57) | **$< 2.2 \times 10^{-16}$** | 1.34 (1.26-1.43) | **$< 2.2 \times 10^{-16}$** |
| **T2D** | 1.63 (1.51-1.76) | **$< 2.2 \times 10^{-16}$** | 1.46 (1.35-1.58) | **$< 2.2 \times 10^{-16}$** | 1.58 (1.41-1.77) | **$3.35 \times 10^{-15}$** | 1.40 (1.24-1.57) | **$2.27 \times 10^{-08}$** |
| **Obesity** | 1.59 (1.48-1.70) | **$< 2.2 \times 10^{-16}$** | 1.52 (1.41-1.62) | **$< 2.2 \times 10^{-16}$** | 1.60 (1.45-1.76) | **$< 2.2 \times 10^{-16}$** | 1.53 (1.39-1.69) | **$< 2.2 \times 10^{-16}$** |
| **CKD** | 1.49 (1.36-1.63) | **$< 2.2 \times 10^{-16}$** | 1.35 (1.23-1.48) | **$4.23 \times 10^{-10}$** | 1.45 (1.27-1.66) | **$< 2.2 \times 10^{-16}$** | 1.27 (1.11-1.45) | **$4.56 \times 10^{-04}$** |
| **Dyslipidaemia** | 1.39 (1.32-1.47) | **$< 2.2 \times 10^{-16}$** | 1.30 (1.23-1.37) | **$< 2.2 \times 10^{-16}$** | 1.41 (1.32-1.52) | **$< 2.2 \times 10^{-16}$** | 1.27 (1.17-1.36) | **$8.78 \times 10^{-10}$** |
| **Any CMC[2]** | 1.49 (1.42-1.55) | **$< 2.2 \times 10^{-16}$** | 1.41 (1.35-1.48) | **$< 2.2 \times 10^{-16}$** | 1.55 (1.46-1.65) | **$< 2.2 \times 10^{-16}$** | 1.43 (1.34-1.52) | **$< 2.2 \times 10^{-16}$** |
| **Any IC plus hypertension** | 1.96 (1.82-2.11) | **$< 2.2 \times 10^{-16}$** | 1.85 (1.72-2.00) | **$< 2.2 \times 10^{-16}$** | 1.87 (1.68-2.08) | **$< 2.2 \times 10^{-16}$** | 1.75 (1.56-1.95) | **$< 2.2 \times 10^{-16}$** |
| **Any IC plus T2D** | 2.25 (1.95-2.58) | **$< 2.2 \times 10^{-16}$** | 2.04 (1.76-2.34) | **$< 2.2 \times 10^{-16}$** | 2.04 (1.64-2.52) | **$8.04 \times 10^{-11}$** | 1.87 (1.50-2.32) | **$1.5 \times 10^{-08}$** |
| **Any IC plus obesity** | 2.15 (1.93-2.38) | **$< 2.2 \times 10^{-16}$** | 2.03 (1.82-2.26) | **$< 2.2 \times 10^{-16}$** | 1.95 (1.65-2.29) | **$1.48 \times 10^{-15}$** | 1.90 (1.61-2.24) | **$2.8 \times 10^{-14}$** |
| **Any IC plus CKD** | 2.05 (1.74-2.39) | **$< 2.2 \times 10^{-16}$** | 1.86 (1.57-2.17) | **$7.85 \times 10^{-14}$** | 2.03 (1.60-2.55) | **$2.81 \times 10^{-09}$** | 1.82 (1.42-2.29) | **$7.9 \times 10^{-07}$** |
| **Any IC plus dyslipidaemia** | 1.87 (1.71-2.04) | **$< 2.2 \times 10^{-16}$** | 1.74 (1.59-1.90) | **$< 2.2 \times 10^{-16}$** | 1.81 (1.59-2.05) | **$< 2.2 \times 10^{-16}$** | 1.66 (1.46-1.89) | **$1.22 \times 10^{-14}$** |
| **Any ICM-MM[3]** | 1.93 (1.81-2.06) | **$< 2.2 \times 10^{-16}$** | 1.83 (1.72-1.96) | **$< 2.2 \times 10^{-16}$** | 1.82 (1.65-1.99) | **$< 2.2 \times 10^{-16}$** | 1.72 (1.56-1.89) | **$< 2.2 \times 10^{-16}$** |

*Supplementary Table 5.* Results of binary logistic regression models comparing the frequency of health outcomes in individuals with physical neglect versus those without. [1]Any IC refers to a diagnosis of depression and/or anxiety from primary or secondary care records (*Supplementary*

*Table 2*). [2]Any CMC refers to a diagnosis of hypertension, T2D, obesity, CKD and/or dyslipidaemia from primary or secondary care records (*Supplementary Table 2*). [3]ICM-MM refers to a diagnosis of any IC plus any CMC from primary or secondary care records. CKD refers to chronic kidney disease (*Supplementary Table 2*). Model 1 is not adjusted for any covariates. Model 2 is adjusted for demographic variables: age, gender, ethnicity and socioeconomic status. Model 3 is adjusted for lifestyle variables: diet, alcohol intake, smoking status and physical activity levels. Model 4 is adjusted for demographic and lifestyle variables: age, gender, ethnicity, socioeconomic status, diet, alcohol intake, smoking status and physical activity levels. All p-values are corrected for multiple testing using the Bonferroni Correction, raw p-values are reported and those formatted in bold survived correction for multiple testing.

| | Emotional Abuse | | | | | | | |
|---|---|---|---|---|---|---|---|---|
| | **Odds ratio (95% CI)** | **p-value** | **Odds ratio (95% CI)** | **p-value** | **Odds ratio (95% CI)** | **p-value** | **Odds ratio (95% CI)** | **p-value** |
| | Model 1<br>Unadjusted | | Model 2<br>Adjusted for demographic variables | | Model 3<br>Adjusted for lifestyle variables | | Model 4<br>Adjusted for demographic and lifestyle variables | |
| **Any IC[1]** | 2.21 (2.12-2.30) | **$< 2.2 \times 10^{-16}$** | 2.04 (1.96-2.13) | **$< 2.2 \times 10^{-16}$** | 2.19 (2.07-2.32) | **$< 2.2 \times 10^{-16}$** | 2.07 (1.95-2.19) | **$< 2.2 \times 10^{-16}$** |
| **Hypertension** | 1.04 (1.00-1.08) | $6.21 \times 10^{-02}$ | 1.30 (1.25-1.35) | **$< 2.2 \times 10^{-16}$** | 1.04 (0.99-1.10) | 0.14 | 1.32 (1.25-1.39) | **$< 2.2 \times 10^{-16}$** |
| **T2D** | 1.31 (1.22-1.40) | **$6.48 \times 10^{-15}$** | 1.50 (1.40-1.61) | **$< 2.2 \times 10^{-16}$** | 1.18 (1.07-1.31) | **$8.43 \times 10^{-04}$** | 1.40 (1.26-1.55) | **$1.02 \times 10^{-10}$** |
| **Obesity** | 1.58 (1.50-1.67) | **$< 2.2 \times 10^{-16}$** | 1.56 (1.47-1.65) | **$< 2.2 \times 10^{-16}$** | 1.58 (1.46-1.71) | **$< 2.2 \times 10^{-16}$** | 1.60 (1.47-1.73) | **$< 2.2 \times 10^{-16}$** |
| **CKD** | 1.12 (1.03-1.21) | $7.87 \times 10^{-03}$ | 1.37 (1.26-1.49) | **$1.06 \times 10^{-13}$** | 1.15 (1.03-1.29) | $1.33 \times 10^{-02}$ | 1.44 (1.28-1.61) | **$9.26 \times 10^{-10}$** |
| **Dyslipidaemia** | 1.09 (1.04-1.13) | **$2.14 \times 10^{-04}$** | 1.34 (1.28-1.40) | **$< 2.2 \times 10^{-16}$** | 1.05 (0.99-1.11) | 0.13 | 1.30 (1.22-1.39) | **$5.96 \times 10^{-16}$** |
| **Any CMC[2]** | 1.12 (1.08-1.15) | **$4.49 \times 10^{-10}$** | 1.38 (1.33-1.43) | **$< 2.2 \times 10^{-16}$** | 1.11 (1.06-1.17) | **$1.03 \times 10^{-05}$** | 1.40 (1.33-1.48) | **$< 2.2 \times 10^{-16}$** |
| **Any IC plus hypertension** | 1.97 (1.85-2.09) | **$< 2.2 \times 10^{-16}$** | 2.07 (1.94-2.20) | **$< 2.2 \times 10^{-16}$** | 1.94 (1.78-2.12) | **$< 2.2 \times 10^{-16}$** | 2.10 (1.92-2.29) | **$< 2.2 \times 10^{-16}$** |
| **Any IC plus T2D** | 2.32 (2.07-2.60) | **$< 2.2 \times 10^{-16}$** | 2.35 (2.09-2.63) | **$< 2.2 \times 10^{-16}$** | 2.15 (1.81-2.55) | **$< 2.2 \times 10^{-16}$** | 2.32 (1.94-2.75) | **$< 2.2 \times 10^{-16}$** |
| **Any IC plus obesity** | 2.64 (2.43-2.87) | **$< 2.2 \times 10^{-16}$** | 2.32 (2.13-2.52) | **$< 2.2 \times 10^{-16}$** | 2.68 (2.38-3.03) | **$< 2.2 \times 10^{-16}$** | 2.47 (2.19-2.79) | **$< 2.2 \times 10^{-16}$** |
| **Any IC plus CKD** | 1.96 (1.71-2.23) | **$< 2.2 \times 10^{-16}$** | 2.14 (1.87-2.45) | **$< 2.2 \times 10^{-16}$** | 1.94 (1.59-2.35) | **$1.73 \times 10^{-11}$** | 2.17 (1.78-2.64) | **$1.2 \times 10^{-14}$** |
| **Any IC plus dyslipidaemia** | 1.94 (1.81-2.08) | **$< 2.2 \times 10^{-16}$** | 2.05 (1.90-2.20) | **$< 2.2 \times 10^{-16}$** | 1.88 (1.69-2.07) | **$< 2.2 \times 10^{-16}$** | 2.02 (1.82-2.24) | **$< 2.2 \times 10^{-16}$** |
| **Any ICM-MM[3]** | 2.08 (1.98-2.19) | **$< 2.2 \times 10^{-16}$** | 2.08 (1.98-2.19) | **$< 2.2 \times 10^{-16}$** | 2.08 (1.93-2.23) | **$< 2.2 \times 10^{-16}$** | 2.14 (1.99-2.31) | **$< 2.2 \times 10^{-16}$** |

*Supplementary Table 6.* Results of binary logistic regression models comparing the frequency of health outcomes in individuals with emotional abuse versus those without. [1]Any IC refers to a diagnosis of depression and/or anxiety from primary or secondary care records (*Supplementary*

*Table 2*). [2]Any CMC refers to a diagnosis of hypertension, T2D, obesity, CKD and/or dyslipidaemia from primary or secondary care records (*Supplementary Table 2*). [3]ICM-MM refers to a diagnosis of any IC plus any CMC from primary or secondary care records. CKD refers to chronic kidney disease (*Supplementary Table 2*). Model 1 is not adjusted for any covariates. Model 2 is adjusted for demographic variables: age, gender, ethnicity and socioeconomic status. Model 3 is adjusted for lifestyle variables: diet, alcohol intake, smoking status and physical activity levels. Model 4 is adjusted for demographic and lifestyle variables: age, gender, ethnicity, socioeconomic status, diet, alcohol intake, smoking status and physical activity levels. All p-values are corrected for multiple testing using the Bonferroni Correction, raw p-values are reported and those formatted in bold survived correction for multiple testing.

| | Physical Abuse | | | | | | | |
|---|---|---|---|---|---|---|---|---|
| | **Odds ratio (95% CI)** | **p-value** | **Odds ratio (95% CI)** | **p-value** | **Odds ratio (95% CI)** | **p-value** | **Odds ratio (95% CI)** | **p-value** |
| | Model 1<br>Unadjusted | | Model 2<br>Adjusted for demographic variables | | Model 3<br>Adjusted for lifestyle variables | | Model 4<br>Adjusted for demographic and lifestyle variables | |
| **Any IC[1]** | 1.80 (1.72-1.88) | **$< 2.2 \times 10^{-16}$** | 1.80 (1.72-1.88) | **$< 2.2 \times 10^{-16}$** | 1.77 (1.66-1.88) | **$< 2.2 \times 10^{-16}$** | 1.77 (1.66-1.89) | **$< 2.2 \times 10^{-16}$** |
| **Hypertension** | 1.15 (1.11-1.20) | **$2.79 \times 10^{-12}$** | 1.32 (1.27-1.38) | **$< 2.2 \times 10^{-16}$** | 1.13 (1.07-1.19) | **$1.44 \times 10^{-05}$** | 1.32 (1.24-1.40) | **$< 2.2 \times 10^{-16}$** |
| **T2D** | 1.54 (1.44-1.65) | **$< 2.2 \times 10^{-16}$** | 1.57 (1.47-1.69) | **$< 2.2 \times 10^{-16}$** | 1.46 (1.32-1.61) | **$1.47 \times 10^{-14}$** | 1.54 (1.39-1.70) | **$< 2.2 \times 10^{-16}$** |
| **Obesity** | 1.74 (1.65-1.84) | **$< 2.2 \times 10^{-16}$** | 1.73 (1.64-1.83) | **$< 2.2 \times 10^{-16}$** | 1.70 (1.56-1.84) | **$< 2.2 \times 10^{-16}$** | 1.72 (1.58-1.86) | **$< 2.2 \times 10^{-16}$** |
| **CKD** | 1.27 (1.17-1.38) | **$1.19 \times 10^{-08}$** | 1.49 (1.36-1.62) | **$< 2.2 \times 10^{-16}$** | 1.29 (1.14-1.44) | **$1.92 \times 10^{-05}$** | 1.50 (1.33-1.69) | **$1.9 \times 10^{-11}$** |
| **Dyslipidaemia** | 1.21 (1.15-1.26) | **$9.42 \times 10^{-16}$** | 1.34 (1.28-1.41) | **$< 2.2 \times 10^{-16}$** | 1.18 (1.10-1.25) | **$4.32 \times 10^{-07}$** | 1.33 (1.25-1.43) | **$< 2.2 \times 10^{-16}$** |
| **Any CMC[2]** | 1.24 (1.20-1.29) | **$< 2.2 \times 10^{-16}$** | 1.42 (1.37-1.48) | **$< 2.2 \times 10^{-16}$** | 1.21 (1.15-1.28) | **$4.91 \times 10^{-14}$** | 1.42 (1.34-1.50) | **$< 2.2 \times 10^{-16}$** |
| **Any IC plus hypertension** | 1.71 (1.60-1.83) | **$< 2.2 \times 10^{-16}$** | 1.82 (1.70-1.94) | **$< 2.2 \times 10^{-16}$** | 1.71 (1.55-1.88) | **$< 2.2 \times 10^{-16}$** | 1.83 (1.66-2.01) | **$< 2.2 \times 10^{-16}$** |
| **Any IC plus T2D** | 2.17 (1.92-2.45) | **$< 2.2 \times 10^{-16}$** | 2.10 (1.86-2.38) | **$< 2.2 \times 10^{-16}$** | 2.08 (1.72-2.48) | **$4.53 \times 10^{-15}$** | 2.09 (1.73-2.51) | **$5.98 \times 10^{-15}$** |
| **Any IC plus obesity** | 2.43 (2.22-2.65) | **$< 2.2 \times 10^{-16}$** | 2.31 (2.11-2.53) | **$< 2.2 \times 10^{-16}$** | 2.40 (2.08-2.71) | **$< 2.2 \times 10^{-16}$** | 2.31 (2.02-2.64) | **$< 2.2 \times 10^{-16}$** |
| **Any IC plus CKD** | 1.88 (1.63-2.17) | **$< 2.2 \times 10^{-16}$** | 2.13 (1.84-2.46) | **$< 2.2 \times 10^{-16}$** | 1.86 (1.50-2.28) | **$4.38 \times 10^{-09}$** | 2.09 (1.69-2.57) | **$5.84 \times 10^{-12}$** |
| **Any IC plus dyslipidaemia** | 1.79 (1.66-1.93) | **$< 2.2 \times 10^{-16}$** | 1.89 (1.75-2.04) | **$< 2.2 \times 10^{-16}$** | 1.78 (1.59-1.98) | **$< 2.2 \times 10^{-16}$** | 1.90 (1.70-2.12) | **$< 2.2 \times 10^{-16}$** |
| **Any ICM-MM[3]** | 1.83 (1.73-1.94) | **$< 2.2 \times 10^{-16}$** | 1.90 (1.80-2.01) | **$< 2.2 \times 10^{-16}$** | 1.81 (1.67-1.96) | **$< 2.2 \times 10^{-16}$** | 1.90 (1.76-2.06) | **$< 2.2 \times 10^{-16}$** |

*Supplementary Table 7.* Results of binary logistic regression models predicting comparing the frequency of health outcomes in individuals with physical abuse versus those without. [1]Any IC refers to a diagnosis of depression and/or anxiety from primary or secondary care records

(*Supplementary Table 2*). [2]Any CMC refers to a diagnosis of hypertension, T2D, obesity, CKD and/or dyslipidaemia from primary or secondary care records (*Supplementary Table 2*). [3]ICM-MM refers to a diagnosis of any IC plus any CMC from primary or secondary care records. CKD refers to chronic kidney disease (*Supplementary Table 2*). Model 1 is not adjusted for any covariates. Model 2 is adjusted for demographic variables: age, gender, ethnicity and socioeconomic status. Model 3 is adjusted for lifestyle variables: diet, alcohol intake, smoking status and physical activity levels. Model 4 is adjusted for demographic and lifestyle variables: age, gender, ethnicity, socioeconomic status, diet, alcohol intake, smoking status and physical activity levels. All p-values are corrected for multiple testing using the Bonferroni Correction, raw p-values are reported and those formatted in bold survived correction for multiple testing.

| | Sexual Abuse | | | | | | | |
|---|---|---|---|---|---|---|---|---|
| | **Odds ratio (95% CI)** | **p-value** | **Odds ratio (95% CI)** | **p-value** | **Odds ratio (95% CI)** | **p-value** | **Odds ratio (95% CI)** | **p-value** |
| | Model 1<br>Unadjusted | | Model 2<br>Adjusted for demographic variables | | Model 3<br>Adjusted for lifestyle variables | | Model 4<br>Adjusted for demographic and lifestyle variables | |
| **Any IC[1]** | 1.61<br>(1.54-1.68) | **$< 2.2 \times 10^{-16}$** | 1.47<br>(1.40-1.54) | **$< 2.2 \times 10^{-16}$** | 1.45<br>(1.37-1.55) | **$< 2.2 \times 10^{-16}$** | 1.36<br>(1.27-1.44) | **$< 2.2 \times 10^{-16}$** |
| **Hypertension** | 0.97<br>(0.93-1.01) | 0.13 | 1.10<br>(1.05-1.14) | **$1.43 \times 10^{-05}$** | 0.97<br>(0.92-1.02) | 0.26 | 1.08<br>(1.02-1.14) | $9.99 \times 10^{-03}$ |
| **T2D** | 1.14<br>(1.06-1.23) | **$4.47 \times 10^{-04}$** | 1.26<br>(1.17-1.36) | **$1.05 \times 10^{-09}$** | 1.16<br>(1.05-1.28) | $3.89 \times 10^{-03}$ | 1.30<br>(1.17-1.44) | **$7.65 \times 10^{-07}$** |
| **Obesity** | 1.43<br>(1.35-1.51) | **$< 2.2 \times 10^{-16}$** | 1.39<br>(1.31-1.48) | **$< 2.2 \times 10^{-16}$** | 1.41<br>(1.30-1.53) | **$5.73 \times 10^{-16}$** | 1.39<br>(1.28-1.51) | **$9.37 \times 10^{-15}$** |
| **CKD** | 1.09<br>(1.00-1.19) | $4.60 \times 10^{-02}$ | 1.19<br>(1.09-1.30) | **$9.11 \times 10^{-05}$** | 1.12<br>(0.99-1.25) | $6.43 \times 10^{-02}$ | 1.22<br>(1.08-1.37) | $8.58 \times 10^{-04}$ |
| **Dyslipidaemia** | 1.02<br>(0.97-1.07) | 0.39 | 1.15<br>(1.10-1.21) | **$8.16 \times 10^{-09}$** | 0.99<br>(0.93-1.05) | 0.74 | 1.10<br>(1.03-1.18) | $2.95 \times 10^{-03}$ |
| **Any CMC[2]** | 1.04<br>(1.01-1.08) | $1.88 \times 10^{-02}$ | 1.17<br>(1.13-1.22) | **$1.14 \times 10^{-15}$** | 1.03<br>(0.98-1.08) | 0.19 | 1.15<br>(1.09-1.21) | **$2.97 \times 10^{-07}$** |
| **Any IC plus hypertension** | 1.49<br>(1.39-1.60) | **$< 2.2 \times 10^{-16}$** | 1.48<br>(1.38-1.58) | **$< 2.2 \times 10^{-16}$** | 1.42<br>(1.29-1.56) | **$9.37 \times 10^{-13}$** | 1.42<br>(1.29-1.57) | **$1.43 \times 10^{-12}$** |
| **Any IC plus T2D** | 1.63<br>(1.42-1.86) | **$5.62 \times 10^{-13}$** | 1.62<br>(1.42-1.85) | **$1.41 \times 10^{-12}$** | 1.65<br>(1.35-1.98) | **$2.08 \times 10^{-07}$** | 1.69<br>(1.39-2.04) | **$8.07 \times 10^{-08}$** |
| **Any IC plus obesity** | 1.97<br>(1.79-2.16) | **$< 2.2 \times 10^{-16}$** | 1.73<br>(1.57-1.90) | **$< 2.2 \times 10^{-16}$** | 1.84<br>(1.60-2.11) | **$< 2.2 \times 10^{-16}$** | 1.69<br>(1.47-1.94) | **$9.94 \times 10^{-14}$** |
| **Any IC plus CKD** | 1.74<br>(1.50-2.00) | **$5.18 \times 10^{-14}$** | 1.73<br>(1.49-2.00) | **$1.58 \times 10^{-13}$** | 1.72<br>(1.39-2.09) | **$2.14 \times 10^{-07}$** | 1.73<br>(1.40-2.12) | **$1.71 \times 10^{-07}$** |
| **Any IC plus dyslipidaemia** | 1.49<br>(1.37-1.61) | **$< 2.2 \times 10^{-16}$** | 1.49<br>(1.37-1.61) | **$< 2.2 \times 10^{-16}$** | 1.41<br>(1.25-1.57) | **$3.11 \times 10^{-09}$** | 1.42<br>(1.26-1.59) | **$2.08 \times 10^{-09}$** |
| **Any ICM-MM[3]** | 1.56<br>(1.47-1.65) | **$< 2.2 \times 10^{-16}$** | 1.49<br>(1.41-1.58) | **$< 2.2 \times 10^{-16}$** | 1.47<br>(1.36-1.59) | **$< 2.2 \times 10^{-16}$** | 1.43<br>(1.32-1.55) | **$5.56 \times 10^{-18}$** |

*Supplementary Table 8.* Results of binary logistic regression models comparing the frequency of health outcomes in individuals with sexual abuse versus those without. [1]Any IC refers to a diagnosis of depression and/or anxiety from primary or secondary care records (*Supplementary Table 2*). [2]Any CMC refers to a diagnosis of hypertension, T2D, obesity, CKD and/or dyslipidaemia from primary or secondary care records (*Supplementary Table 2*). [3]ICM-MM refers to a diagnosis of any IC plus any CMC from primary or secondary care records. CKD refers to chronic kidney disease (*Supplementary Table 2*). Model 1 is not adjusted for any covariates. Model 2 is adjusted for demographic variables: age, gender, ethnicity and socioeconomic status. Model 3 is adjusted for lifestyle variables: diet, alcohol intake, smoking status and physical activity levels. Model 4 is adjusted for demographic and lifestyle variables: age, gender, ethnicity, socioeconomic status, diet, alcohol intake, smoking status and physical activity levels. All p-values are corrected for multiple testing using the Bonferroni Correction, raw p-values are reported and those formatted in bold survived correction for multiple testing.

| Health outcomes | Emotional Neglect Odds ratio[4] (Females) (95% CI) | Odds ratio[5] (Males) (95% CI) | p-value of difference[6] |
|---|---|---|---|
| **Any IC[1]** | 1.87 (1.78-1.98) | 1.77 (1.65-1.90) | 0.29 |
| **Hypertension** | 1.20 (1.13-1.26) | 1.09 (1.03-1.15) | 0.05 |
| **T2D** | 1.36 (1.21-1.52) | 1.16 (1.06-1.27) | 0.04 |
| **Obesity** | 1.41 (1.29-1.52) | 1.24 (1.13-1.35) | 0.03 |
| **CKD** | 1.19 (1.05-1.33) | 1.008 (0.90-1.13) | 0.07 |
| **Dyslipidaemia** | 1.26 (1.16-1.32) | 1.08 (1.02-1.14) | $5.14 \times 10^{-03}$ |
| **Any CMC[2]** | 1.28 (1.22-1.34) | 1.11 (1.05-1.16) | **$1.25 \times 10^{-04}$** |
| **Any IC plus hypertension** | 1.80 (1.65-1.97) | 1.61 (1.45-1.78) | 0.14 |
| **Any IC plus T2D** | 1.92 (1.57-2.35) | 1.79 (1.48-2.15) | 0.57 |
| **Any IC plus obesity** | 1.92 (1.69-2.17) | 1.81 (1.52-2.15) | 0.55 |
| **Any IC plus CKD** | 1.78 (1.46-2.17) | 1.36 (1.05-1.75) | 0.12 |
| **Any IC plus dyslipidaemia** | 1.85 (1.67-2.06) | 1.61 (1.43-1.81) | 0.10 |
| **Any ICM-MM[3]** | 1.87 (1.74-2.01) | 1.66 (1.52-1.81) | 0.05 |

*Supplementary Table 9.* Results of binary logistic regression models comparing the frequency of health outcomes in individuals with emotional neglect versus those without in females and males separately. [1]Any IC refers to a diagnosis of depression and/or anxiety from primary or secondary care records (*Supplementary Table 2*). [2]Any CMC refers to a diagnosis of hypertension, T2D, obesity, CKD and/or dyslipidaemia from primary or secondary care records (*Supplementary Table 2*). [3]ICM-MM refers to a diagnosis of any IC plus any CMC from primary or secondary care records. CKD refers to chronic kidney disease (*Supplementary Table 2*). [4,5]Odds ratios and confidence intervals, adjusted for age, ethnicity, socioeconomic status, diet, alcohol intake, smoking status and physical activity levels, are reported for females and males separately. [6]P-values of the interaction between ACEs and gender, adjusted for age, ethnicity, socioeconomic status, diet, alcohol intake, smoking status and physical activity levels, are reported. All p-values are corrected for multiple testing using the Bonferroni Correction, raw p-values are reported and none survived correction for multiple testing.

| Health outcomes | Physical Neglect | | |
|---|---|---|---|
| | Odds ratio[4] (Females) (95% CI) | Odds ratio[5] (Males) (95% CI) | p-value of difference[6] |
| **Any IC[1]** | 1.60 (1.45-1.76) | 1.56 (1.37-1.77) | 0.83 |
| **Hypertension** | 1.34 (1.22-1.47) | 1.34 (1.22-1.47) | 0.98 |
| **T2D** | 1.44 (1.19-1.72) | 1.35 (1.16-1.57) | 0.35 |
| **Obesity** | 1.65 (1.44-1.88) | 1.37 (1.17-1.60) | 0.07 |
| **CKD** | 1.40 (1.16-1.68) | 1.13 (0.93-1.37) | 0.20 |
| **Dyslipidaemia** | 1.32 (1.18-1.47) | 1.21 (1.09-1.35) | 0.19 |
| **Any CMC[2]** | 1.44 (1.32-1.57) | 1.40 (1.27-1.54) | 0.67 |
| **Any IC plus hypertension** | 1.78 (1.54-2.06) | 1.69 (1.42-2.00) | 0.60 |
| **Any IC plus T2D** | 1.66 (1.19-2.26) | 2.06 (1.51-2.74) | 0.69 |
| **Any IC plus obesity** | 2.05 (1.67-2.48) | 1.57 (1.14-2.12) | 0.16 |
| **Any IC plus CKD** | 2.14 (1.59-2.82) | 1.29 (0.81-1.94) | 0.09 |
| **Any IC plus dyslipidaemia** | 1.78 (1.50-2.10) | 1.51 (1.23-1.84) | 0.14 |
| **Any ICM-MM[3]** | 1.79 (1.58-2.01) | 1.61 (1.38-1.87) | 0.27 |

*Supplementary Table 10.* Results of binary logistic regression models comparing the frequency of health outcomes in individuals with physical neglect versus those without in females and males separately. [1]Any IC refers to a diagnosis of depression and/or anxiety from primary or secondary care records (*Supplementary Table 2*). [2]Any CMC refers to a diagnosis of hypertension, T2D, obesity, CKD and/or dyslipidaemia from primary or secondary care records (*Supplementary Table 2*). [3]ICM-MM refers to a diagnosis of any IC plus any CMC from primary or secondary care records. CKD refers to chronic kidney disease (*Supplementary Table 2*). [4,5]Odds ratios and confidence intervals, adjusted for age, ethnicity, socioeconomic status, diet, alcohol intake, smoking status and physical activity levels, are reported for females and males separately. [6]P-values of the interaction between ACEs and gender, adjusted for age, ethnicity, socioeconomic status, diet, alcohol intake, smoking status and physical activity levels, are reported. All p-values are corrected for multiple testing using the Bonferroni Correction, raw p-values are reported and none survived correction for multiple testing.

| Health outcomes | Emotional Abuse | | |
|---|---|---|---|
| | Odds ratio[4] (Females) (95% CI) | Odds ratio[5] (Males) (95% CI) | p-value of difference[6] |
| **Any IC[1]** | 1.98 (1.85-2.12) | 2.28 (2.06-2.52) | 9.85 x $10^{-03}$ |
| **Hypertension** | 1.36 (1.27-1.47) | 1.27 (1.16-1.39) | 0.61 |
| **T2D** | 1.25 (1.07-1.46) | 1.54 (1.34-1.76) | 0.03 |
| **Obesity** | 1.59 (1.43-1.76) | 1.60 (1.40-1.82) | 0.89 |
| **CKD** | 1.57 (1.35-1.82) | 1.27 (1.05-1.52) | 0.09 |
| **Dyslipidaemia** | 1.36 (1.25-1.48) | 1.25 (1.14-1.38) | 0.45 |
| **Any CMC[2]** | 1.43 (1.34-1.53) | 1.37 (1.26-1.49) | 0.60 |
| **Any IC plus hypertension** | 2.04 (1.83-2.28) | 2.22 (1.92-2.56) | 0.19 |
| **Any IC plus T2D** | 1.81 (1.39-2.32) | 2.94 (2.30-3.71) | 7.85 x $10^{-03}$ |
| **Any IC plus obesity** | 2.30 (1.98-2.66) | 2.91 (2.32-3.61) | 0.12 |
| **Any IC plus CKD** | 2.20 (1.72-2.78) | 2.15 (1.49-2.99) | 0.91 |
| **Any IC plus dyslipidaemia** | 1.84 (1.61-2.10) | 2.34 (1.99-2.75) | 8.68 x $10^{-03}$ |
| **Any ICM-MM[3]** | 2.05 (1.87-2.25) | 2.34 (2.06-2.64) | 0.05 |

*Supplementary Table 11.* Results of binary logistic regression models comparing the frequency of health outcomes in individuals with emotional abuse versus those without in females and males separately. [1]Any IC refers to a diagnosis of depression and/or anxiety from primary or secondary care records (*Supplementary Table 2*). [2]Any CMC refers to a diagnosis of hypertension, T2D, obesity, CKD and/or dyslipidaemia from primary or secondary care records (*Supplementary Table 2*). [3]ICM-MM refers to a diagnosis of any IC plus any CMC from primary or secondary care records. CKD refers to chronic kidney disease (*Supplementary Table 2*). [4,5]Odds ratios and confidence intervals, adjusted for age, ethnicity, socioeconomic status, diet, alcohol intake, smoking status and physical activity levels, are reported for females and males separately. [6]P-values of the interaction between ACEs and gender, adjusted for age, ethnicity, socioeconomic status, diet, alcohol intake, smoking status and physical activity levels, are reported. All p-values are corrected for multiple testing using the Bonferroni Correction, raw p-values are reported and none survived correction for multiple testing.

| Health outcomes | Physical Abuse | | |
|---|---|---|---|
| | Odds ratio[4] (Females) (95% CI) | Odds ratio[5] (Males) (95% CI) | p-value of difference[6] |
| **Any IC[1]** | 1.81 (1.67-1.97) | 1.71 (1.54-1.89) | 0.41 |
| **Hypertension** | 1.37 (1.26-1.49) | 1.27 (1.17-1.38) | 0.57 |
| **T2D** | 1.49 (1.25-1.76) | 1.56 (1.37-1.76) | 0.61 |
| **Obesity** | 1.82 (1.63-2.04) | 1.58 (1.40-1.78) | 0.07 |
| **CKD** | 1.63 (1.37-1.93) | 1.39 (1.18-1.64) | 0.24 |
| **Dyslipidaemia** | 1.36 (1.23-1.51) | 1.31 (1.20-1.43) | 0.95 |
| **Any CMC[2]** | 1.49 (1.38-1.60) | 1.35 (1.25-1.46) | 0.16 |
| **Any IC plus hypertension** | 1.98 (1.74-2.25) | 1.65 (1.42-1.90) | 0.14 |
| **Any IC plus T2D** | 1.91 (1.43-2.51) | 2.21 (1.71-2.81) | 0.57 |
| **Any IC plus obesity** | 2.17 (1.83-2.56) | 2.54 (2.03-3.14) | 0.34 |
| **Any IC plus CKD** | 2.25 (1.70-2.93) | 1.87 (1.32-2.58) | 0.58 |
| **Any IC plus dyslipidaemia** | 1.94 (1.66-2.26) | 1.85 (1.57-2.16) | 0.87 |
| **Any ICM-MM[3]** | 1.99 (1.79-2.20) | 1.79 (1.58-2.03) | 0.34 |

*Supplementary Table 12.* Results of binary logistic regression models comparing the frequency of health outcomes in individuals with physical abuse versus those without in females and males separately. [1]Any IC refers to a diagnosis of depression and/or anxiety from primary or secondary care records (*Supplementary Table 2*). [2]Any CMC refers to a diagnosis of hypertension, T2D, obesity, CKD and/or dyslipidaemia from primary or secondary care records (*Supplementary Table 2*). [3]ICM-MM refers to a diagnosis of any IC plus any CMC from primary or secondary care records. CKD refers to chronic kidney disease (*Supplementary Table 2*). [4,5]Odds ratios and confidence intervals, adjusted for age, ethnicity, socioeconomic status, diet, alcohol intake, smoking status and physical activity levels, are reported for females and males separately. [6]P-values of the interaction between ACEs and gender, adjusted for age, ethnicity, socioeconomic status, diet, alcohol intake, smoking status and physical activity levels, are reported. All p-values are corrected for multiple testing using the Bonferroni Correction, raw p-values are reported and none survived correction for multiple testing.

| Health outcomes | Sexual Abuse | | |
|---|---|---|---|
| | **Odds ratio[4] (Females) (95% CI)** | **Odds ratio[5] (Males) (95% CI)** | **p-value of difference[6]** |
| **Any IC[1]** | 1.36 (1.26-1.46) | 1.35 (1.19-1.52) | 0.96 |
| **Hypertension** | 1.12 (1.04-1.20) | 1.03 (0.95-1.13) | 0.34 |
| **T2D** | 1.41 (1.21-1.63) | 1.21 (1.04-1.40) | 0.21 |
| **Obesity** | 1.45 (1.31-1.61) | 1.29 (1.12-1.48) | 0.17 |
| **CKD** | 1.14 (0.97-1.34) | 1.34 (1.12-1.59) | 0.16 |
| **Dyslipidaemia** | 1.13 (1.04-1.24) | 1.08 (0.98-1.19) | 0.63 |
| **Any CMC[2]** | 1.18 (1.11-1.26) | 1.09 (1.002-1.19) | 0.22 |
| **Any IC plus hypertension** | 1.41 (1.24-1.58) | 1.47 (1.25-1.73) | 0.57 |
| **Any IC plus T2D** | 1.67 (1.28-2.14) | 1.71 (1.27-2.27) | 0.97 |
| **Any IC plus obesity** | 1.64 (1.39-1.92) | 1.82 (1.39-2.36) | 0.56 |
| **Any IC plus CKD** | 1.63 (1.25-2.09) | 2.01 (1.40-2.81) | 0.29 |
| **Any IC plus dyslipidaemia** | 1.40 (1.21-1.61) | 1.47 (1.22-1.77) | 0.64 |
| **Any ICM-MM[3]** | 1.43 (1.30-1.58) | 1.44 (1.24-1.67) | 0.89 |

*Supplementary Table 13.* Results of binary logistic regression models comparing the frequency of health outcomes in individuals with sexual abuse versus those without in females and males separately. [1]Any IC refers to a diagnosis of depression and/or anxiety from primary or secondary care records (*Supplementary Table 2*). [2]Any CMC refers to a diagnosis of hypertension, T2D, obesity, CKD and/or dyslipidaemia from primary or secondary care records (*Supplementary Table 2*). [3]ICM-MM refers to a diagnosis of any IC plus any CMC from primary or secondary care records. CKD refers to chronic kidney disease (*Supplementary Table 2*). [4,5]Odds ratios and confidence intervals, adjusted for age, ethnicity, socioeconomic status, diet, alcohol intake, smoking status and physical activity levels, are reported for females and males separately. [6]P-values of the interaction between ACEs and gender, adjusted for age, ethnicity, socioeconomic status, diet, alcohol intake, smoking status and physical activity levels, are reported. All p-values are corrected for multiple testing using the Bonferroni Correction, raw p-values are reported and none survived correction for multiple testing.

**Supplementary references**


Byrne, N. M., Hills, A. P., Hunter, G. R., Weinsier, R. L. and Schutz, Y. 2005. Metabolic equivalent: one size does not fit all. *Journal of Applied Physiology* 99, pp. 1112-1119. Available at: https://journals.physiology.org/doi/pdf/10.1152/japplphysiol.00023.2004

Daviet, R. et al. 2022. Associations between alcohol consumption and gray and white matter volumes in the UK Biobank. *Nature communications* 13(1), p. 1175. Available at: https://pmc.ncbi.nlm.nih.gov/articles/PMC8897479/pdf/41467_2022_Article_28735.pdf

Hepsomali, P. and Groeger, J. A. 2021. Diet and general cognitive ability in the UK Biobank dataset. *Scientific reports* 11(1), p. 11786. Available at: https://pmc.ncbi.nlm.nih.gov/articles/PMC8175590/pdf/41598_2021_Article_91259.pdf

Liu, W., Wang, T., Zhu, M. and Jin, G. 2023. Healthy diet, polygenic risk score, and upper gastrointestinal cancer risk: a prospective study from UK Biobank. *Nutrients* 15(6), p. 1344. Available at: https://mdpi-res.com/d_attachment/nutrients/nutrients-15-01344/article_deploy/nutrients-15-01344.pdf?version=1678440287

Wang, M., Zhou, T., Song, Q., Ma, H., Hu, Y., Heianza, Y. and Qi, L. 2022. Ambient air pollution, healthy diet and vegetable intakes, and mortality: a prospective UK Biobank study. *International journal of epidemiology* 51(4), pp. 1243-1253. Available at: https://pmc.ncbi.nlm.nih.gov/articles/PMC9365625/pdf/dyac022.pdf
